# Genomic network analysis characterizes genetic architecture and identifies trait-specific biology

**DOI:** 10.1101/2024.12.03.24318432

**Authors:** Jackson G. Thorp, Zachary F. Gerring, William R. Reay, Eske M. Derks, Andrew D. Grotzinger

## Abstract

Pervasive genetic overlap across human complex traits necessitates developing multivariate methods that can parse pleiotropic and trait-specific genetic signals. Here, we introduce Genomic Network Analysis (GNA), an analytic framework that applies the principles of network modelling to estimates of genetic overlap derived from genome-wide association study (GWAS) summary statistics. The result is a genomic network that describes the conditionally independent genetic associations between traits that remain when controlling for shared signal with the broader network of traits. Graph theory metrics provide added insight by formally quantifying the most important traits in the genomic network. GNA can discover additional trait-specific pathways by incorporating gene expression or genetic variants into the network to estimate their conditional associations with each trait. Extensive simulations establish GNA is well-powered for most GWAS. Application to a diverse set of traits demonstrate that GNA yields critical insight into the genetic architecture that demarcate genetically overlapping traits at varying levels of biological granularity.

## Introduction

Pleiotropic genetic variants associated with multiple traits are the norm rather than the exception^1^, with only 10% of variants estimated to be trait-specific^2^. Widespread pleiotropy will, in part, reflect the fact that most traits under study are the multifaceted consequence of core disease pathways and secondary risk factors that are both being captured in GWAS estimates. Comprehensive characterization of GWAS findings thereby necessitates parsing shared and unique signal across sets of genetically correlated traits. Refinement of more specific genetic signals would assist to prioritise cross-trait genetic overlaps and GWAS loci that should be subjected to further experimental or clinical interrogation. For example, complex human diseases often show pervasive genetic overlap with circulating biochemical markers like lipids and enzymes^3,4^; however, it is difficult to understand which estimates of genetic overlap represent true shared biology as opposed to what arises due to pleiotropy between these biochemical traits. In response to these challenges, we introduce Genomic Network Analysis (GNA), a flexible framework for network modelling of multivariate GWAS data, enabling the identification of conditional genetic associations at genome-wide, genetic variant, and gene centric levels of analysis.

Network modelling has been used to describe complex multivariate relationships within a wide range of biological, social, economic and psychological systems (e.g., gene co-expression networks, neural networks, social networks, psychopathological networks)^5^. Rather than focus on the functioning of individual system components, network approaches examine how these components interact and are organised within the larger system. A graphical model, for example, reflects a network of conditionally independent relationships (edges) between a set of variables (nodes). Once a network model is estimated, properties of the network such as topological features, sparsity, node centrality, and node clustering, can provide important insight into the structure and functioning of the overall system^6^.

Leveraging these principles of network models in the context of genetics with GNA allows for the characterization of genetic signals that arise only due to associations with other variables in the network as compared to those that are specific to a particular trait. At the genome-wide level, this can take a complex web of genetic correlations between a set of traits and deconvolve to a sparse network that describes the most critical connections. The construction of genome-wide networks in GNA requires only GWAS summary statistics as input and can be applied to GWAS data from participant samples with varying and unknown degrees of sample overlap. GNA can also incorporate into the network individual genetic variants (network GWAS) or individual genes (network TWAS), facilitating the identification of trait-specific biology by estimating of effect of a variant or gene conditional on other traits in the network. Below we validate GNA via extensive simulations that demonstrate this approach is well-calibrated, adequately powered, and has high accuracy for sample sizes and SNP-based heritability estimates typical of modern day GWAS. Through a range of empirical applications to real-world GWAS, we demonstrate GNA’s ability to provide novel insight into disease aetiology and the identification of trait-specific disease pathways that are otherwise masked by overlapping signal with genetic correlates.

## Results

### Overview of GNA

GNA estimates a network model for a set of traits at a genomic level, where nodes represent the genetic component of each trait (ℎ*^g^*), which are connected by edges representing the partial genetic correlation (*Pr_g_*) between traits (i.e. the genetic correlation between a pair of traits while conditioning on all other traits in the network). Specifically, our approach fits a Gaussian graphical model (GGM)^7^ to a genetic variance-covariance matrix (obtained from multivariable LDSC^8,9^). Therefore, GNA requires only GWAS summary statistics as input and can be applied to GWAS data from participant samples with varying and unknown degrees of participant sample overlap.

GNA implements a stepdown model search procedure to select a sparse network model; a saturated (fully connected) GGM is estimated; non-significant parameters are pruned and the GGM is refit recursively until only significant edges remain in the network. This edge selection process serves to produce a network that reflects a more parsimonious and readily interpretable system of relationships. We provide several model fit statistics to assess the adequacy of the sparse network model in describing the data. Such model fit statistics are typically calculated using participant sample (*N*). However, our genomic network approach allows for including GWAS summary statistics that will often have varying sample sizes. To circumvent this issue, we develop and validate via simulations a summary-based approach for calculating common model fit metrics such as comparative fix index (CFI)^10^ and standardized root mean square residual (SRMR)^11^.

### Simulations

Simulations were conducted to evaluate GNA across a realistic range of population generating scenarios that varied the three key parameters related to power in the genomic network: sample size, number of traits in the network (K), and SNP-based heritability (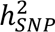). We find that GNA has well-controlled Type I error across all simulating conditions (Fig 1A), and produces unbiased parameter estimates (Fig 1C and 1D) and well-calibrated standard errors in the presence of participant sample overlap (ratio between estimated standard errors and standard deviation of point estimates = 1.01). The simulations suggest that GNA has sufficient statistical power (>0.80) to estimate a 5 trait network when trait 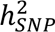 Z > 7, a 10 trait network when 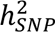 Z > 17, a 15 trait network when 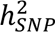 Z > 21, and a 20 trait network when 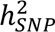 Z > 23 (Fig 1B).

**Figure 1:**
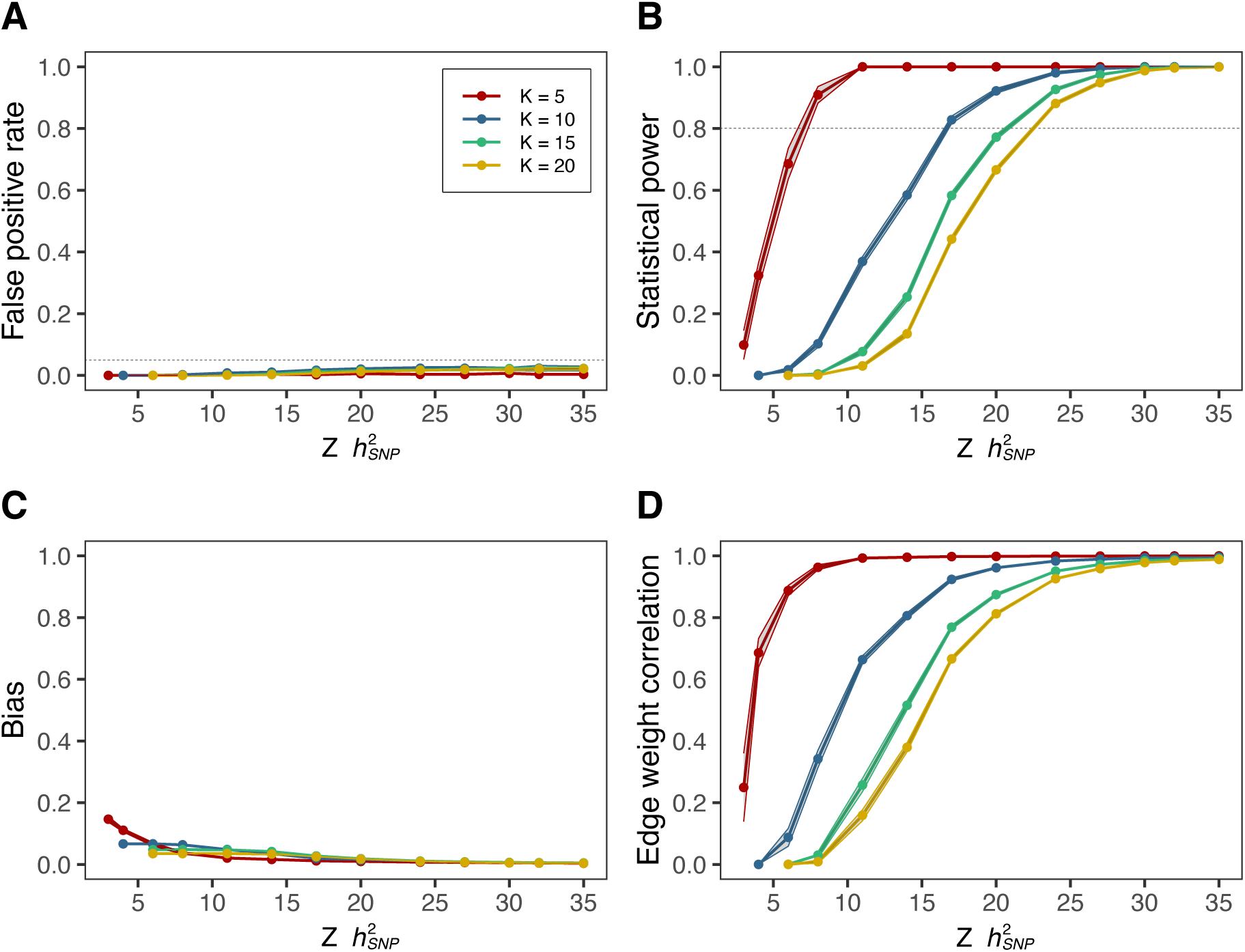
GNA simulation results. Results from 12,000 simulation runs of GNA varying the number of traits in the network (K) and the power of traits in the network (SNP-based heritability Z statistic (Z ℎ^$^); which is dependent upon both the SNP-based heritability and sample size of the traits). Each dot represents the mean value across all simulation runs for that set of parameters, with 95% confidence intervals plotted as bands. **A)** False positive rate, where a ‘positive’ represents a non-zero edge weight and a ‘negative’ represents a null edge weight. **B)** Statistical power (false negative rate). **C)** Bias in estimated edge weights, calculated as the mean of the absolute differences between the true network edge weights and estimated edge weights. **D)** Correlation between the set of estimated edge weights and the set of true edge weights within each run.

Criticisms of phenotypic networks include poor reliability^12^ or low accuracy^13^ that stem from estimating partial correlations with small participant sample sizes. By comparison, GWAS data often has extremely large participant sample sizes to ensure adequate power for genetic discovery. Importantly, we find that, given the thresholds above, we have high accuracy, as indicated by the percentage of 95% confidence intervals that include the population generating parameter and high reliability (indexed by the fact that the estimated edges are correlated at or near 1 with the population generating edges).

### Conditional genetic relationships between type 2 diabetes and cardiometabolic traits

First, we demonstrate the simplest use of GNA – to identify conditional genetic relationships (partial genetic correlations) between a set of traits. We applied GNA to East Asian ancestry GWAS of type 2 diabetes (T2D) and a set of cardiometabolic traits that have all been previously observationally and genetically implicated to be involved in the pathogenesis of T2D^14,15^, including fasting plasma glucose (FPG), body mass index (BMI), systolic blood pressure (SBP), triglycerides (TG) and high-density lipoprotein cholesterol (HDL; **Suppl. Table 1**). T2D and the five measured cardiometabolic traits are all significantly genetic correlated with each other (Fig 2A).

**Figure 2.**
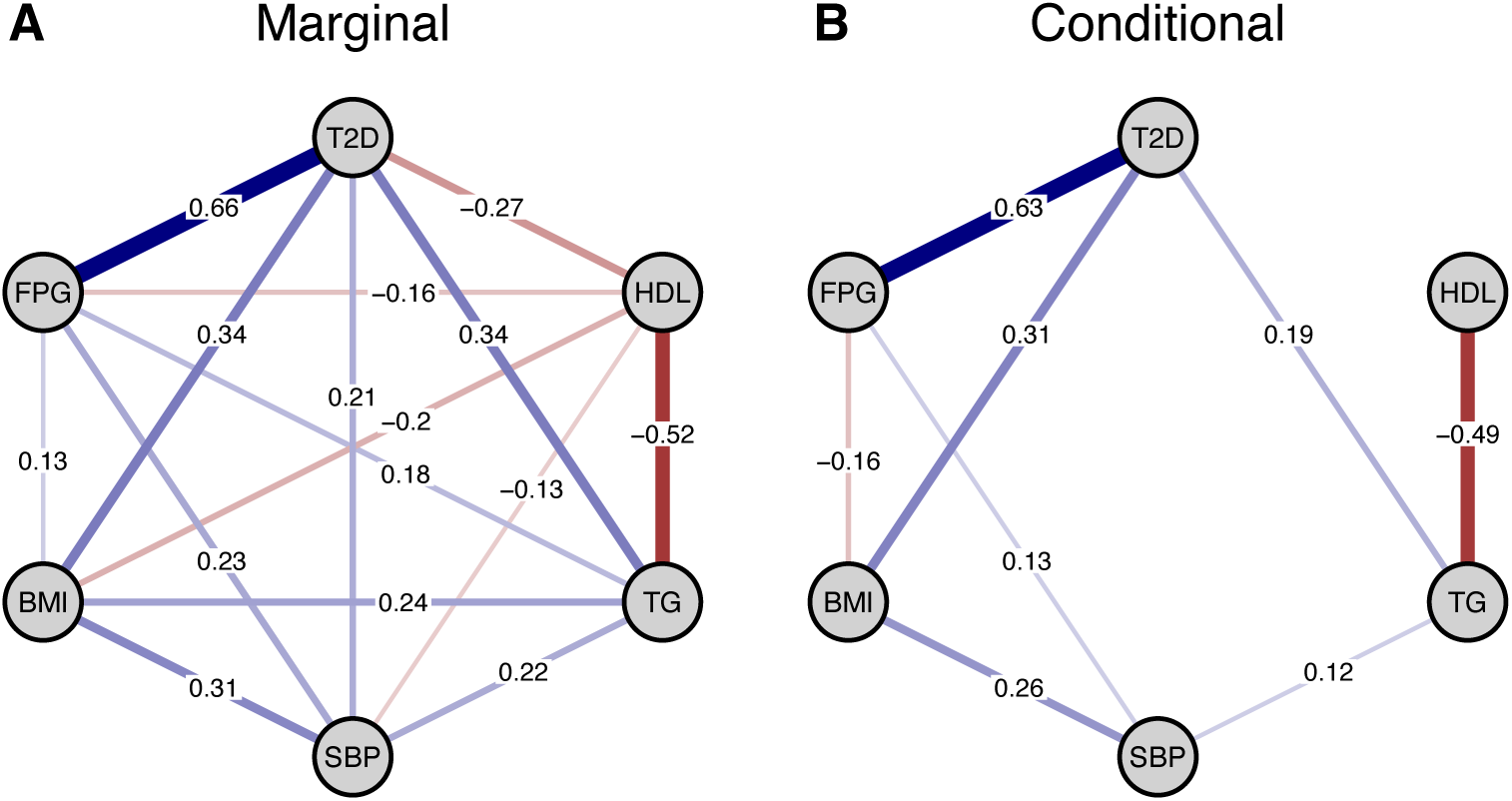
Network of type 2 diabetes (T2D) and other cardiometabolic traits. **A)** Genetic correlations (marginal associations) between T2D and cardiometabolic traits estimated via LD Score Regression. **B)** Partial genetic correlations (conditional associations) between T2D and cardiometabolic traits estimated via GNA after non-significant parameters (FDR > 0.05) are pruned from the network. T2D = Type 2 Diabetes; FPG = fasting plasma glucose; BMI = body mass index; SBP = systolic blood pressure; TG = triglycerides; HDL = high-density lipoprotein cholesterol.

We applied GNA to these traits, which identified 8 significant partial genetic correlations. In other words, from the 15 total trait pair associations, only 8 remained significant after conditioning on all other traits (Fig 2B, edge density = 0.53). To assess the extent to which these 8 partial correlations capture the full genetic overlap between the traits, we estimated a sparse network model in GNA, in which non-significant parameters are fixed to zero and the remaining network parameters are re-estimated (Fig 2B; **Suppl. Table 2**). The network model had excellent fit to the observed genetic covariance matrix (CFI = 0.97, SRMR = 0.04), implying the sparse network captures the genetic overlap between the cardiometabolic traits well.

We highlight two broad types of conditional relationships that emerged from this analysis. As expected, the strong genetic overlap between T2D and FPG remained very consistent in the sparse network conditioned on all remaining traits (*r_g_* = 0.66, se = 0.05, p = 6.9×10^-43^; *Pr*_%_= 0.65, se = 0.04, p = 6.9×10^-75^), with similar findings for BMI (*r_g_* = 0.34, se = 0.03, p = 6.7×10^-29^; *Pr_g_*= 0.29, se = 0.04, p = 2.7×10^-15^). In contrast, T2D and HDL levels were conditionally independent, such that their association (*r_g_* = -0.27, se = 0.03, p = 1.1×10^-19^) did not hold given the other traits in the network (*Pr*_%_= -0.06, se = 0.05, p = 0.20). This finding is in line with previous genetic and observational evidence that has challenged the existence of direct relationship between lower circulating HDL and liability to T2D in a similar vein to what has been found with respect to HDL and cardiovascular disease^16–18^.

### Genetic Network Analysis of Neuroticism Items Reveals Crucial Components

Phenotypic networks are increasingly applied in the psychiatric literature to gain clinically relevant insight into the symptoms that most likely affect disease onset and progression^19,20^. How important an item is in a network can be formally quantified using developed graph theory metrics that are included as part of the output from the GNA R package. Here we show how two of these metrics, expected influence and clustering coefficients (**Online Supplement** and **Suppl. Figure 1** for additional metrics), can be used to identify critical items within a genomic network of 12 neuroticism items from the Eysenck Personality Questionnaire Revised-Short Form^21^ (**Suppl. Table 3; Method**). Expected influence is calculated as the sum of the edge weights connected to a given node^22^, thereby quantifying the direct effects of a focal node on the remaining nodes. Clustering coefficients assess the degree to which an individual node’s neighbours (the other nodes it is connected to) are interconnected. A high clustering coefficient of a given node indicates that its neighbours are highly connected. These coefficients have been discussed as a measure of the node’s redundancy in the network, where removing a node with a larger clustering coefficient would still produce a similar network^23^. The combination of high centrality values and low clustering coefficients can demarcate key nodes in a network.

LDSC estimated genetic correlations for 12 neuroticism items in the UK Biobank^21^ were all highly significant (maximum *p*-value = 4.36 x 10^-22^) and revealed pervasive genetic overlap (mean *r_g_* = .63; range = .37 - .89). The full set of genetic correlations, and even partial genetic correlations, form a complex web of interconnections that are difficult to interpret (Fig 2A). The application of GNA, which recursively estimates the network model until only significant edges remain, produced a much sparser and more interpretable network that continued to provide good fit to the data (CFI = .98; SRMR = .08; Fig 2B; **Suppl. Table 4**). Evaluating node importance in this sparse network revealed that nervousness had the largest expected influence and, relative to other nodes in the network, a lower clustering coefficient (**Suppl. Table 5**). This indicates that the genetic signal for nervousness reflects a crucial component that uniquely links various aspects of neuroticism. As neuroticism is a robust predictor of a range of adverse outcomes, including risk for psychopathology^23^, life satisfaction^26^, and mortality^27^, this may reflect a more efficacious intervention target within this predictive personality construct.

### Genomic Network Analysis of Biochemical Markers

We then applied GNA to 19 blood biochemical traits from the UK Biobank^4^ (**Suppl. Table 6**). From the 171 total trait pairs, 115 were significantly genetically correlated (edge density = 0.67). We estimated a sparse genomic network model in GNA, which reduced this set of cross-trait correlations to 37 (Fig 4A, edge density = 0.22, **Suppl. Table 7**). The model had good fit to the data (CFI = .920; SRMR = .055) and recapitulated strong genetic relationships between biochemical traits that are known to be biologically related – for example, large genetic overlaps remained in the sparse network between enzymes used clinically as markers of liver function (gamma-glutamyl transferase, alanine aminotransferase, and aspartate aminotransferase).

**Figure 3.**
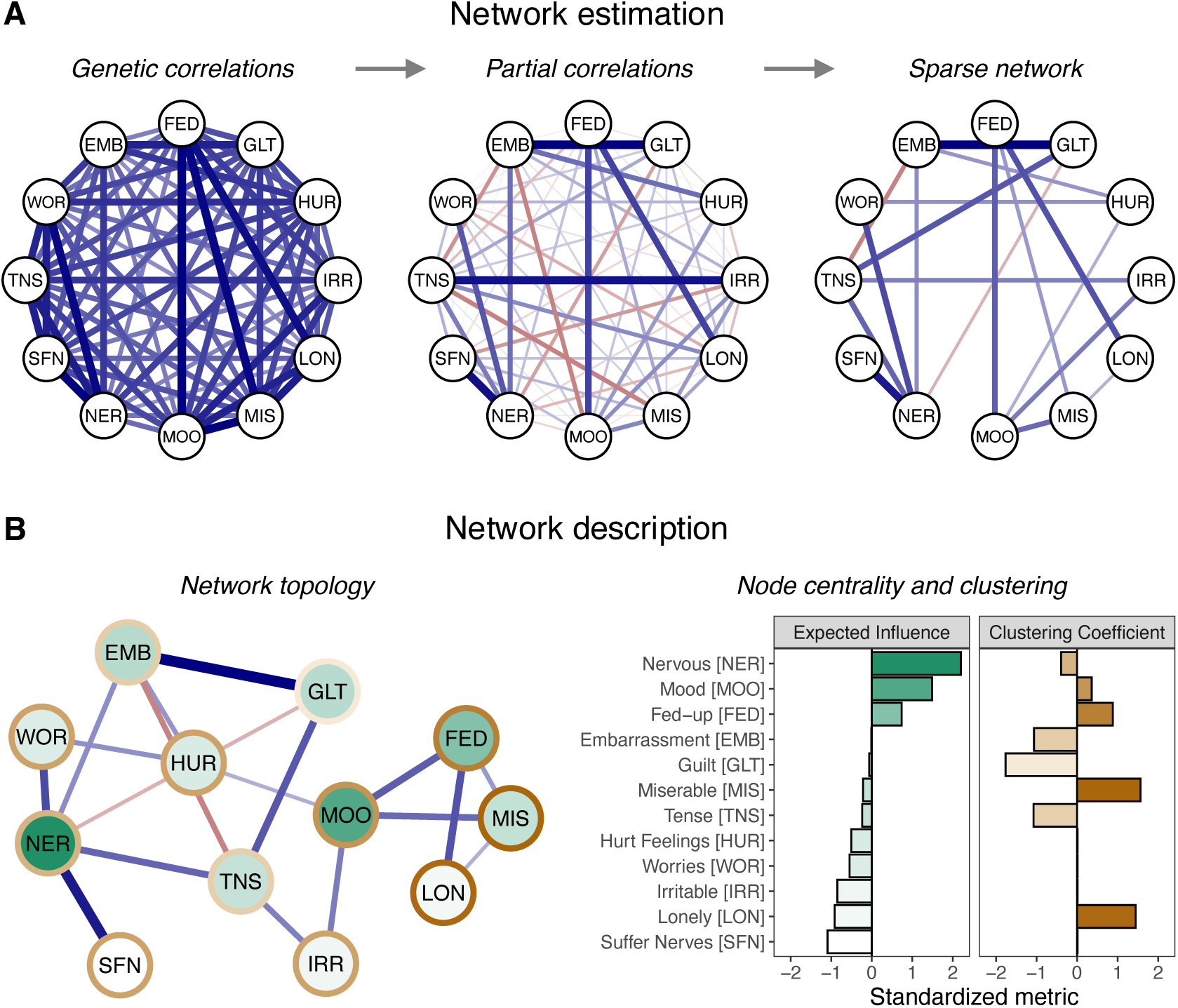
Neuroticism Network Results. **A)** Genetic correlations between the 12 neuroticism items, the full set of partial genetic correlations, and the pruned, sparse network model (FDR < 0.05). Edge thickness corresponds to the size of the edge weight, with blue and red edges indicating positive and negative values, respectively. **B)** Left: the neuroticism network plotted using multidimensional scaling. The nodes are shaded according to their centrality in the network and the borders of the nodes are shaded according to their clustering coefficient. Right: expected influence (measure of node centrality) and the Zhang clustering coefficient^24^ for each node in the network, standardized relative to all other nodes in the network.

**Figure 4.**
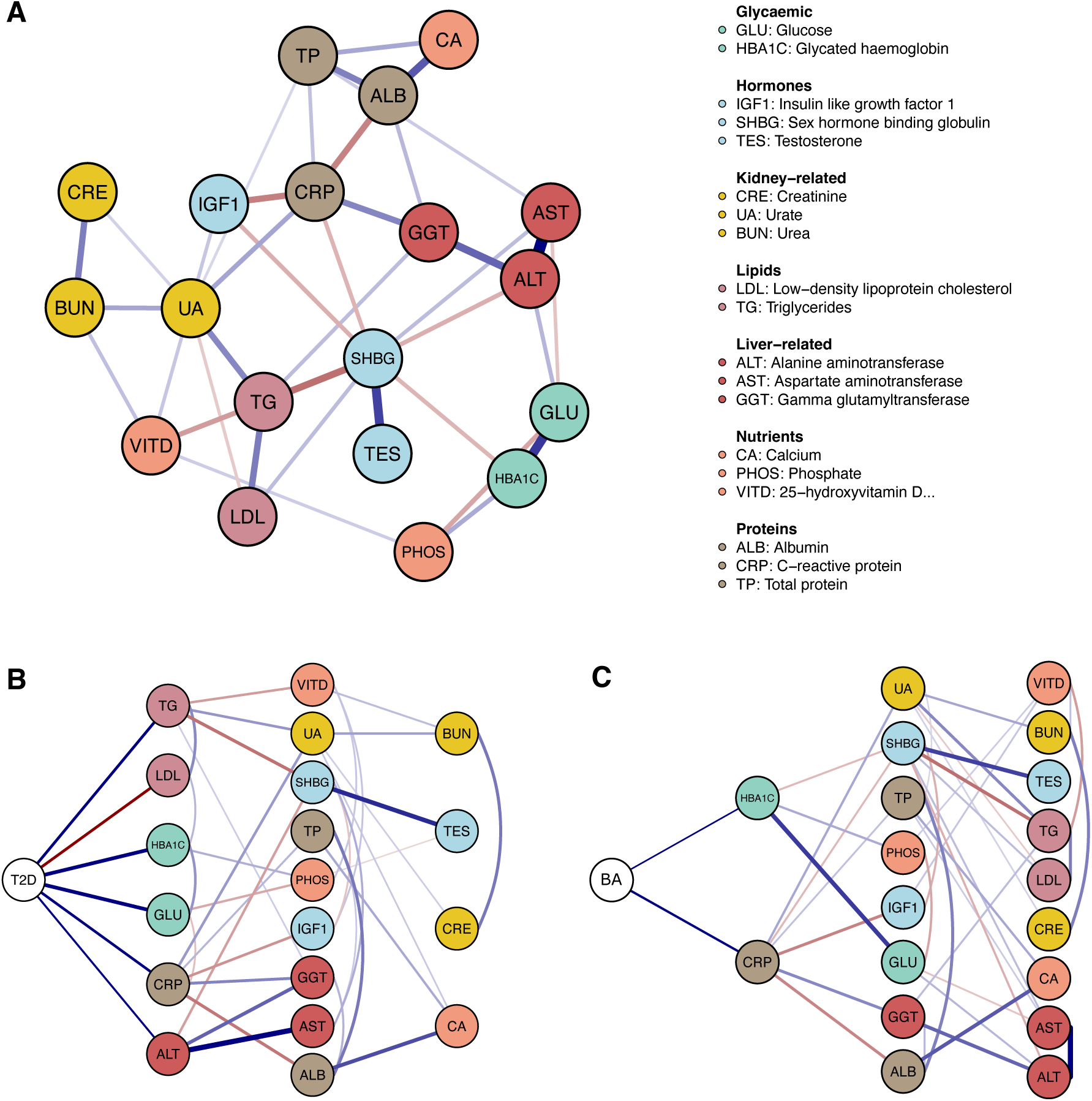
Genomic network of biochemical markers. **A)** Genomic network of 19 biochemical markers from the UK Biobank. Sparse network model estimated via GNA by recursively pruning non-significant edges (*P*bonf < 0.05) from the network. Edge thickness corresponds to the size of the edge weight, with blue and red edges indicating positive and negative values, respectively. **B)** Conditional genetic associations (partial genetic correlations) between type 2 diabetes (T2D) and the biochemical markers, estimated via a sparse network model in GNA. **C)** Conditional genetic associations between bronchial asthma (BA) and the biochemical markers.

Once a network is estimated, insight into its structure can be gained by assessing the global properties of the network (that is, the properties of the overall network rather than of any individual nodes). GNA provides three main metrics of global network structure: the global clustering coefficient (degree of clustering across the whole network)^28^, average path length (average distance between nodes in the network)^29^, and the small-worldness index (the degree to which the network possesses small-world properties, i.e. high clustering and a short average path length)^30,31^. The small-worldness index of the biochemical trait network was 0.42, suggesting its structure lies somewhere between a random and a small-world network. Further inspection of the metrics show that the average path length (2.19) was extremely similar to that from equivalent random networks (mean = 2.14; 2.5% and 97.5% quantiles = [1.89,2.31]), and the clustering coefficient (0.25) was less than found in equivalent lattice networks (mean = 0.45; 2.5% and 97.5% quantiles = [0.35,0.54]). This suggests that while the biochemical trait network has a short average path length, it does not possess a high degree of clustering, and therefore does not show a clear small-world structure.

Next, we incorporated disease traits into the biochemical network as a hypothesis free search for relevant and conditionally associated disease-biochemical marker genetic overlap. First, we included T2D, which was genetically correlated with 14 of the 19 biomarkers. However, application of GNA revealed that, after controlling for the other biomarkers, T2D was only associated with 6 biomarkers (**Suppl. Table 8)**. In line with glycaemic dysregulation being the key pathological process of T2D, glycated haemoglobin (HbA1c; *Pr_g_*= 0.38, se = 0.033) and glucose (random blood sample, GLU; *Pr_g_*= 0.39, se = 0.042) still exhibited the strongest correlation estimates, followed by triglycerides (TG; *Pr_g_*= 0.26, se = 0.040), low-density lipoprotein cholesterol (LDL; *Pr_g_*= -0.25, se = 0.041), C-reactive protein (CRP; *Pr*_%_= 0.25, se = 0.040), and alanine transaminase (ALT; *Pr_g_*= 0.16, se = 0.036). We note that the preserved negative genetic correlation between LDL and T2D is of interest given the putative relationship between statin usage and increased risk of T2D^32^. Further, our data aligns with existing genetic evidence, such as the inverse relationship between the key LDL-associated gene *PCSK9* with T2D relative to coronary artery disease that may arise due to the role of *PCSK9* in pancreatic islets^33,34^, although further work is needed to fully understand the role of LDL in T2D pathogenesis.

Second, we analysed cross-trait genetic relationships between the respiratory disorder asthma and the 19 biochemical measures in the same fashion, with asthma liability also genetically correlated with 14 of the 19 biomarkers. The network revealed that asthma was conditionally associated with just 2 biomarkers, including C-reactive protein (CRP, *Pr_g_*= 0.20, se = 0.027) and glycated haemoglobin (HbA1c, *Pr_g_*= 0.11, se = 0.024; **Suppl. Table 9**). The conditional positive correlation between asthma and CRP, which is used as an inflammatory biomarker, is biologically plausible, whilst potential shared biology with HbA1c may arise due to the impact of elevated blood glucose on lung function and the immune system^35,36^.

### Network GWAS in GNA

#### Overview

A GWAS for any given trait reflects a mixture of the biological pathways that are most proximal to that trait, along with the pleiotropic pathways shared with genetic correlates. Methodologies that can separate out these trait-specific and pleiotropic pathways offer the opportunity to better characterize and understand emergent GWAS signal. To this end, GNA can incorporate individual genetic variants (SNPs) into the network to produce GWAS summary statistics that reflect the conditional associations between the SNP and each trait in the network. This can provide insight into the biological pathways that are unique to, or more strongly associated with, a given trait. In an empirical application using the same East Asian genetic ancestry summary statistics for T2D and related metabolic traits presented above we show how network GWAS can uncover primary disease pathways that are otherwise masked by overlapping signal with genetically correlated outcomes.

#### Network GWAS reveals core Type 2 Diabetes disease pathways

We focus here on findings for T2D, but a list of significantly associated loci and corresponding Manhattan plots are provided for the other traits in the network in **Suppl. Tables 10-15** and **Suppl. Figures 2-6.** Fifty significant loci (*p* < 5 x 10^-8^) were conditionally associated with the unique genetic variance for T2D (**Method** for definition of locus). **Figure 5** depicts a genetic variant (rs12507026) that was genome-wide significant for the univariate GWAS of BMI (*p* = 1.02 x 10^-19^) and T2D (*p* = 1.72 x 10^-8^). Results from the network GWAS revealed this same variant was also associated with the genetic variance unique to BMI (*p* = 5.02 x 10^-10^) but was not conditionally associated with T2D (*p* = 0.83). This variant falls within the gene region for *THAP12P9* and *PRDX4P1* and has been previously linked to childhood obesity^37^ and BMI^38^ in external European samples.

**Figure 5:**
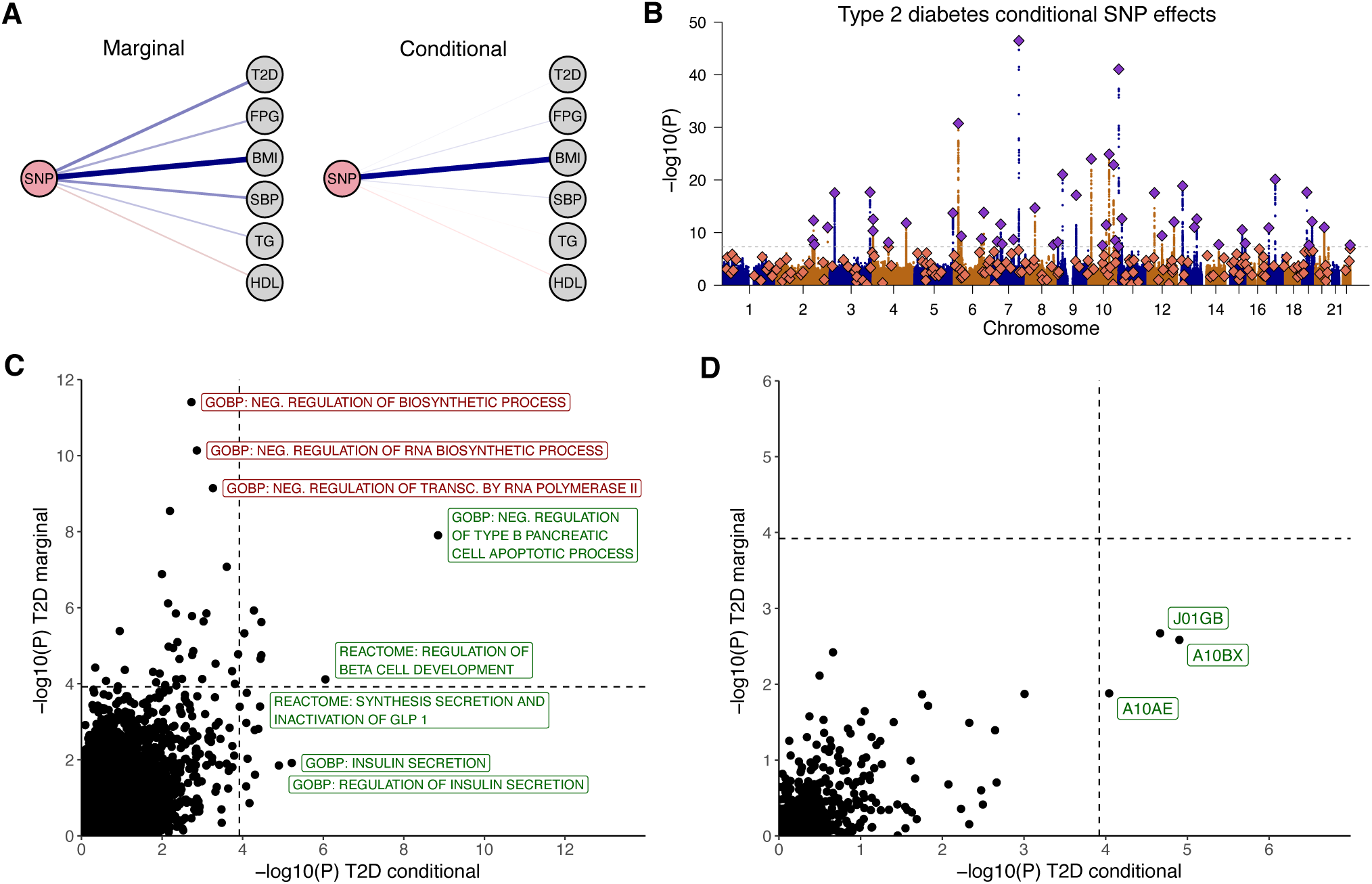
Network GWAS of type 2 diabetes and cardiometabolic traits. **A)** Associations between a genetic variant (rs12507026) and the included traits from univariate GWAS (marginal) and network GWAS (conditional). The width of the line indicates the effect size. While this variant initially had broad associations across traits only the conditional association with BMI remained significant in the network GWAS. **B)** Manhattan plot of conditional genetic associations for T2D from the network GWAS. Independent significant loci that were significant for both network GWAS and univariate GWAS results are depicted as purple diamonds, whereas loci that were only significant for univariate GWAS are shown as orange diamonds. The grey dashed line denotes the genome-wide significance threshold (*p* < 5e-8). The final two panels depict scatter plots of gene set enrichment results for **C**) gene sets from the Molecular Signatures Database and **D**) gene sets defined by Anatomical Therapeutic Chemical (ATC) codes, obtained when using the conditional T2D results from network GWAS (x-axis) and univariate GWAS (y-axis) as input. Grey dashed lines in both panels denote the p-value at which FDR < 0.05. Specific results depicted in green were significant using the network GWAS results whereas gene sets annotated in red reflect those that were significant only when using the univariate GWAS results.

The signal captured by univariate GWAS and network GWAS results for T2D were characterized using gene set analyses (GSA) performed in MAGMA^39^. GSA results estimated using the univariate GWAS effects reflected extremely broad biological pathways (**Suppl. Table 16**), including the top result (*negative regulation of biosynthetic processes*, *p* = 3.93 x 10^-12^) for a gene set capturing biological processes that inhibit chemical reactions in the body. There were 16 novel gene sets that were identified using the network GWAS, but not univariate GWAS, results as input. Within these novel gene sets, the most significant reflected the biological processes that define T2D^40^, namely insulin secretion (*p* = 6.04 x 10^-6^) and pancreatic β-cell pathways (*p* = 3.74 x 10^-5^; **Suppl. Table 17**). The third most significant novel gene set, glucagon like peptide 1 (GLP-1) pathways (*p* = 3.71 x 10^-5^), also reflects the biology targeted by GLP-1 receptor agonists that have demonstrated clinical efficacy in T2D management^41^. Further, we performed GSA using gene sets defined by Anatomical Therapeutic Chemical (ATC) codes from DrugBank^42^. The conditional T2D GWAS effects were significantly enriched in three ATC code gene sets: A10BX (*Other blood glucose lowering drugs, excl. insulins*; *p* = 1.25 x 10^-5^), A10AE (*Insulins and analogues for injection, long-acting*; *p* = 9.06 x 10^-5^), and J01GB (*Other aminoglycosides*; *p* = 2.15 x 10^-5^). Notably, two of these codes indicate drugs that are currently used to treat diabetes. In contrast, the univariate T2D GWAS effects were not significantly enriched in any ATC code gene-set (**Suppl. Table 18**). This illustrates how the GNA framework can remove broadly pleiotropic signal with genetic correlates to produce conditional SNP-level associations that provide unique insight into disease specific biology.

### Network TWAS in GNA

#### Overview

Gene expression data in disease relevant tissue types (e.g., specific brain regions) can be costly and difficult to obtain. TWAS circumvents these pragmatic barriers by imputing gene expression using genetic variants associated with expression and integrating their effect size with that of those same variants from GWAS summary statistics for a trait of interest. These functional weights are publicly available for most tissues and are calculated from secondary, reference datasets that include both genotypes and gene expression data. The network TWAS extension within GNA uses this TWAS output to incorporate imputed gene expression into the genomic network (**Method; Online Supplement**). As in other GNA applications, estimated edges between a gene and trait in the network reflect partial genetic associations controlling for genetic overlap with the other nodes in the network. These conditional associations capture the conditional effect of gene expression on each trait, thereby providing insight into trait-specific biology. We highlight the utility of network TWAS through application to a set of psychiatric disorders.

#### Characterizing Disorder-specific Gene Expression across Psychiatric Disorders

Phenotypic comorbidity^43^ and genetic overlap^44^ are pervasive across psychiatric disorders, raising central questions about etiological divergence. We began by fitting a genome-wide network model to 7 well-powered psychiatric disorders (i.e., SNP-based heritability Z-statistic > 10, which was deemed appropriate based on reported simulations above; **Suppl. Table 19**). These traits were schizophrenia (SCZ)^45^, bipolar disorder (BIP)^46^, major depressive disorder (MDD)^47^, autism spectrum disorder (ASD)^48^, cannabis use disorder (CUD)^49^, anorexia nervosa (AN)^50^, and attention-deficit hyperactivity disorder (ADHD)^51^. The network structure at the genome-wide level fit the data well (CFI = .99, SRMR = .03; **Figure 6; Suppl. Table 20**). The strongest partial correlation in the broader network was observed between bipolar disorder and schizophrenia (*pr_g_* = .60, *SE* = .03), with the next largest between ADHD and CUD (*pr_g_* = .57, *SE* = .07), followed by a conditional association between two neurodevelopmental disorders (ADHD and ASD; *pr_g_* = .46, *SE* = .07).

**Figure 6.**
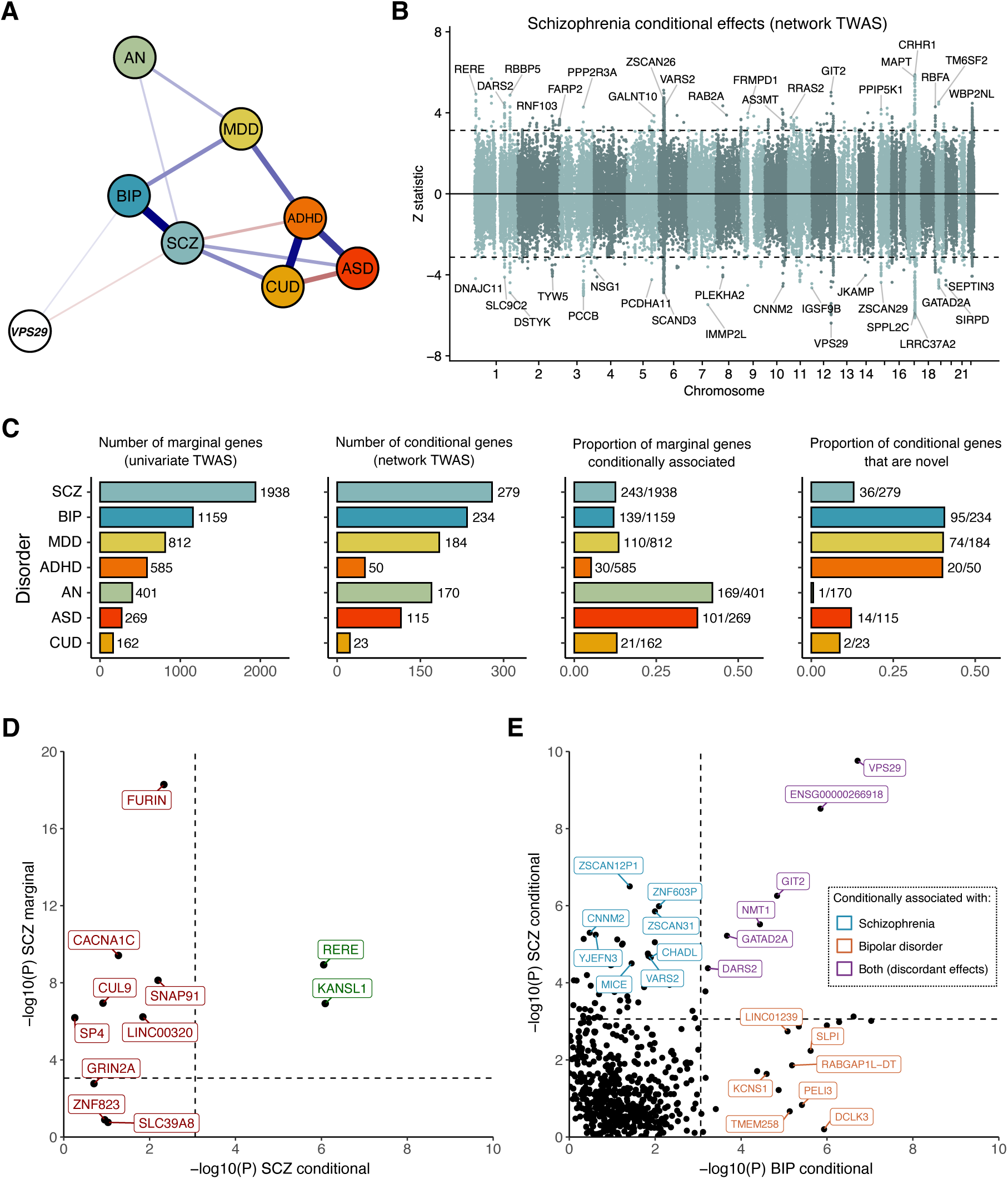
Network TWAS of psychiatric disorders. **A**) The psychiatric network model incorporating gene expression effects for the gene *VPS29,* that was estimated to have directionally opposing (antagonistic) effects on schizophrenia and bipolar disorder in the network TWAS. **B**) Miami plot of network TWAS results for schizophrenia. The dotted horizontal lines depicts the threshold for significance (FDR < 0.05). **C)** Number of genes significantly associated (FDR < 0.05) with each disorder in the univariate TWAS (marginal genes); number of genes significantly associated (FDR < 0.05) with each disorder in the network TWAS (conditional genes); proportion of genes associated with each disorder in the univariate TWAS that remained significantly associated in the network TWAS; proportion of conditional genes that were novel relative to the marginal genes (i.e. genes associated with each disorder in the network TWAS that were not significantly associated in the univariate GWAS). **D)** univariate TWAS vs. network TWAS results for 11 high confidence genes prioritised for schizophrenia. **E)** Schizophrenia vs. bipolar disorder conditional associations for genes that were associated with both disorders in univariate TWAS. *AN:* anorexia nervosa; *BIP:* bipolar disorder; *MDD:* major depressive disorder; *CUD:* cannabis use disorder*; ADHD:* attention-deficit/hyperactivity disorder.

Univariate imputed gene expression was calculated using the FUSION TWAS package^52^ in combination with functional weights from 14 public brain-tissue datasets from GTEx v8^53^ and PsychENCODE^54^. These models of imputed gene expression (genetically predicted expression) for each psychiatric disorder was then added to the genomic network model to run the network TWAS, which estimated the association between each gene and the residual (disorder-specific) genetic variance for each psychiatric disorder not accounted for by the broader network (Figure 6A, **Suppl. Tables 21-25** for full set of results across disorders).

Firstly, we discuss the findings for SCZ. The network TWAS identified 279 genes whose imputed expression was significantly associated with the residual genetic variance in SCZ not shared with the other disorders in the genomic network (Figure 6B,C). We highlight how modelling these conditional associations with SCZ can refine known risk genes that may have a more specific relationship with SCZ liability, as opposed to more pleiotropic, transdiagnostic effects in psychiatry. To explore this, the univariate versus network TWAS results for the most confidently prioritised SCZ risk genes from the most recent psychiatric genomics consortium SCZ GWAS^45^ were extracted from our results (Figure 6D, Methods, N = 11 with TWAS data available). This revealed that two of these high-confidence SCZ risk genes (*RERE* and *KANSL1*, both involved in epigenetic regulation) remained conditionally associated with schizophrenia and are functionally interesting in the context of SCZ. For instance, *RERE* is a gene that is known to be involved in the regulation of the genomic actions retinoic acid via its canonical nuclear receptor repertoire^55^, with altered retinoic signalling previously been linked to SCZ through multiple-lines of evidence, including preliminary positive clinical trial outcomes, as reviewed previously^56^. Conversely, the remaining high-confidence genes were not significant upon performing the network modelling across the different psychiatric disorders, suggesting that they may have more pleiotropic, transdiagnostic relevance.

Secondly, modelling conditional associations between genetically predicted expression and liability to each psychiatric disorder using this network approach also revealed several novel risk genes (FDR < 0.05) that did not survive multiple-testing correction in the univariate analyses (Figure 6C). We provide these novel signals uncovered as conditionally associated with each of the disorders as a resource to the literature to further characterise their biological significance, with two examples presented forthwith. In ADHD, the most conditionally significant novel finding was with genetically predicted expression of the gene *IRAK2* (*Z* = -4.07, *p* = 4.66 x 10^-5^, PsychENCODE cortical panel), which is a kinase that is involved in signal transduction after the interleukin-1 receptor is stimulated^57^. *IRAK2* was only nominally associated with ADHD in the univariate TWAS (*p* = 2.23 x 10^-3^), demonstrating how modelling conditional relationships between the psychiatric disorders can reveal novel insights. We were also able to uncover novel genes for CUD using this approach, which is one of the psychiatric disorders with less known risk genes to date. For example, the most significant novel conditionally significant gene associated with CUD was the gene that encodes Adipocyte Enhancer-Binding Protein 1 (*AEBP1*, *Z* = 3.60, *p* = 3.18 x 10-4, *q* = 0.03, Basal ganglia panel), a gene known to be involved in processes including adipogenesis and pro-inflammatory signalling^58^.

Finally, we highlight how genes for which genetically predicted expression was associated with multiple disorders, but with opposite directions of effect, can be recovered by our network approach (Figure 6A, 6E). Upon comparing the conditionally significant signals uncovered for SCZ and BIP, respectively, we found genes like *VPS29* as strong conditional signals for both disorders but with discordant direction of effect. For example, increased genetically predicted expression of *VPS29* in the anterior cingulate cortex was associated with increased odds of SCZ (*p* = 6.22 x 10^-8^, *q* = 3.23 x 10^-5^), whilst increased genetically predicted expression in that same brain region was found to be protective for BIP (*p* = 1.94 x 10^-7^, *q* = 8.67 x 10^-5^). Given that the *VPS29* is a purported to have important neuronal functionality^59^, further mechanistic dissection of its differential relationship with two disorders is warranted in light of their considerable overlap in clinical features and genetic architecture. However, we do emphasise that further finemapping of this region would be needed to confirm that the same underlying causal variants contribute to both SCZ and BIP in the *VPS29* region. In summary, our application of genetically predicted expression to a sparse, cross-disorder psychiatric network both refined more specific association signals, as well as uncovering novel putative risk genes.

## Discussion

Here we introduce Genomic Network Analysis (GNA), an analytic tool that applies the principles of network analysis to model the conditional genetic associations across multivariate systems of traits. GNA can take an otherwise dense structure of genetic correlations and deconvolve it to a sparse genomic network that retains the most crucial components. The sparse network can be further characterized using centrality metrics, clustering coefficients, global metrics, and graphing options made available through the open-source GNA R package. Our empirical results demonstrate how tangible insights into the most critical items in a network of neuroticism and relevant associations between biomarkers and clinical outcomes can be gained though its application. As GNA takes GWAS summary statistics as input, it can also move beyond phenotypic approaches to incorporate rare or even mutually exclusive traits with varying and unknown levels of participant sample overlap within the same statistical model.

GNA additionally offers the ability to incorporate other biological units of analysis into the genomic network. Via an empirical application to seven psychiatric disorders, we show how GNA can be used to identify patterns of gene expression that biologically differentiate the traits in the network. At the level of individual genetic variants, we apply GNA to pinpoint SNPs that have conditionally significant associations with T2D and its clinical correlates. Follow-up analyses revealed that standard univariate GWAS captured broad biological pathways (e.g., downward regulation of chemical reactions). This was compared to network GWAS results that identified novel biological pathways relevant to the clinically defining features (i.e., insulin secretion; pancreatic β-cells), treatment targets (e.g., GLP-1 pathways) and existing interventions (e.g., glucose-lowering drugs) for T2D. This demonstrates how GNA can refine genetic signal to pull out core trait biology that is otherwise masked by the broadly pleiotropic pathways captured by traditional univariate GWAS. While the biology for T2D is well-defined, future applications of GNA stand to provide valuable insights into the fundamental biological processes that delineate disease states with currently ambiguous etiologies.

GNA has unique advantages relative to existing approaches. Network analysis is often juxtaposed with factor analysis, a multivariate approach used to model shared architecture via latent factors, which can be implemented for GWAS data using Genomic SEM^9^. Relative to factor models, networks do not require an a priori specification of structure and are uniquely identified^2^. In addition, while factor models are well-suited for identifying shared risk pathways, GNA reflects a useful tool for identifying unique signal via its estimation of conditional genetic associations. Among existing approaches that are focused on trait-specific signal (e.g. mtCOJO^60^, GWAS-by-subtraction^61^, LAVA^62^), GNA stands apart from these methods with respect to its ability to examine networks, and trait-specific signal therein, in an integrated framework that can incorporate multiple levels of biological analysis. Even with these distinctions, multiple methods in human complex trait genomics designed to assess trait-specific genetic pathways carries the advantage of being able to produce particularly robust lines of evidence via triangulation across methods^5^.

The GNA framework has several limitations. As the estimates from LDSC are used as input to the genomic network, any biases from LDSC will transfer over to GNA. In particular, we highlight recent work showing that cross-trait assortative mating (xAM) can bias estimates of genetic overlap from GWAS data^63^, though also note that many genetic correlations are sizeable enough that xAM is highly unlikely to be the sole driver of these estimates and phenotypes like biochemical traits with simpler genetic architectures are less likely to be impacted by upward bias due to xAM^64^. Recent whole-genome sequencing efforts indicate rare variants harbor a substantial proportion of trait heritability^65^. As the current GNA framework is limited to common genetic variants, results should be interpreted as capturing this specific component of the genetic signal. An exciting avenue for future methods development reflects extending recent work detailing genetic correlations estimated from rare variants^66^ to multivariate frameworks like GNA. We note that when the traits in the network are theorized to have causal links that the GNA network GWAS framework could be applied in future work as a multivariate tool for identifying genetic instruments for Mendelian randomization analyses. This is because SNPs that are initially associated with multiple traits, but are only conditionally associated with a primary trait in the network, satisfy the exclusion restriction assumption^67^ and are in line with a model of vertical pleiotropy where the SNP only affects the secondary traits via its association with the primary trait.

A standard GWAS will capture a mixture of biological pathways that are more directly associated with the phenotype along with pleiotropic pathways shared with genetic correlates. This is evident in the fact that the vast majority of associated genetic variants are likely pleiotropic^2^ and sizeable genetic correlations are observed across even disparate phenotypes^8^. Here we validate and empirically apply GNA, a multivariate genomic framework that uses the principles of network analysis to parse these shared and trait-specific pathways. GNA can thereby produce more detailed etiological models by identifying and characterizing the biological boundaries that genetically distinguish correlated outcomes at the genome-wide, gene expression, and genetic variant level of analysis.

## Methods

### Gaussian graphical model

A network represents a system of conditionally independent relationships (edges) between a set of variables (nodes). In the case of a Gaussian graphical model (GGM)^7^, the degree of conditional independence between two variables is represented by partial correlation coefficients – the correlation between two variables that results after partialling out all other variables in the network. The partial correlations between a set of variables *Y* are computed by standardising elements of the inverse of the variance-covariance matrix Σ:

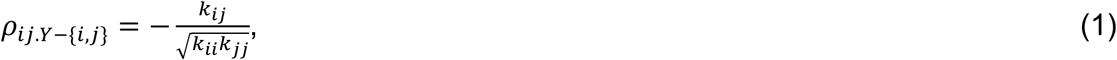

where *κ* is an element of the precision matrix, *K* = Σ*^0^, and *Y* − {*i*, *j*} denotes the set of variables excluding variable *i* and *j*. These partial correlation coefficients are directly used to represent edge weights in the network; if the partial correlation coefficient between two variables is zero, there is conditional independence and hence no edge in the network.

The model-implied variance-covariance matrix 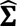 of a GGM can be formed as:

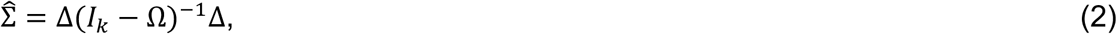

where Ω is a symmetric weight matrix with zeros on the diagonal and off-diagonal element ω*_ij_*_’_ represents the edge weight between node *i* and node *j*, Δ is a diagonal scaling matrix which is a function of the diagonal of the precision matrix 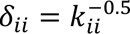, and *I_k_*> is an identity matrix of size *k*^68^. Given that sample partial correlations will almost never be exactly zero even when the population partial correlation is zero, the partial correlation matrix represents a fully connected network, where all nodes are connected to all other nodes (i.e. the model is saturated; 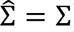). A sparse network can be obtained by constraining some elements of Ω to be zero (i.e. assuming that the two nodes are conditionally independent given all other nodes in the network).

### Network model estimation

GNA takes an empirical genetic variance-covariance matrix (*S*) and its associated sampling covariance matrix (*V_s_*; obtained from multivariable LDSC^8,9^) to estimate a GGM. The *S* matrix reflects the SNP-based heritabilities (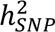) on the diagonal and genetic covariances on the off-diagonal (genetic correlations [*r_g_*] in the standardized case). The *V_s_* matrix has as many columns as there are unique elements in the *S* matrix and is estimated directly from the GWAS data using a block jackknife resampling procedure. The diagonal of *V* contains the sampling variances (the squared standard errors [*SEs*]) for each of the estimates populating the *S* matrix, which allow for GWAS with varying precision to be included in the same model. The off-diagonal contains sampling dependencies, which allow for GWAS with varying and unknown levels of participant sample overlap to be appropriately modelled. GNA uses the *psychonetrics* R package for specification of the GGM and numerical optimization^69^.

Network parameters are obtained in GNA using a maximum likelihood (ML) estimator, which minimises the following fit function:

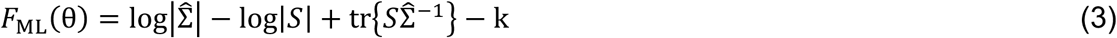

To obtain accurate *SEs* of model parameters we apply a sandwich correction to the sampling covariance matrix:

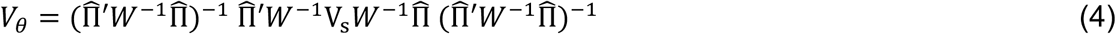

Where 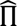 is the Jacobian matrix of model derivatives evaluated at the parameter estimates, 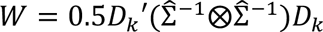., and *V_s_* is the sampling covariance matrix of Σ obtained using multivariable LDSC. Specifically, 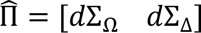, where *d*Σ_8_ and *d*Σ_Δ_ are the derivatives of Σ with respect to Ω and Δ, respectively:

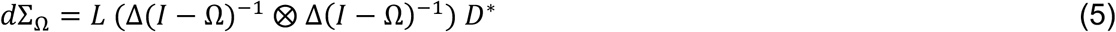

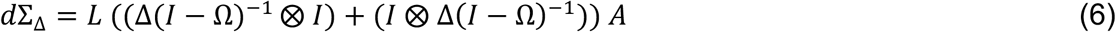

*L* is an elimination matrix, *I* is an identity matrix, *D\** is a duplication matrix, and *A* is a diagonalisation matrix.

### Edge selection

A key benefit of network analysis is the ability to take a complex web of relationships and distil it down to a more tractable, sparse network. This can be achieved in GNA by pruning non-significant edges at some significance threshold. The default behaviour in the GNA R package is for this edge pruning to occur via an iterative process. This process begins with estimating the full network of partial genetic correlations, followed by pruning out all non-significant edges, re-estimating the network with those non-significant edges fixed to 0 (i.e., specified to be conditionally independent), and repeating this network estimation process until only significant edges remain. An alternative to pruning on significance described in the phenotypic network literature is to control the network sparsity using some form of regularisation technique (e.g., a graphical lasso [glasso]^70^). We do not employ this approach for several reasons. First, regularisation methods rely on a single participant sample size. As one of the benefits of GNA reflects the ability to incorporate GWAS of varying size there is not a single sample size that can be used to describe the genomic network. Second, simulation studies have shown that glasso regularization produces more estimation errors at larger sample sizes^71^. As GWAS datasets are by necessity much larger than typical phenotypic data, this suggests that regularization techniques may not be well suited for these kinds of genomic dataset. In addition, regularization approaches assume minimal true connections in the population (i.e., a highly sparse population generating network)^72^. Given pervasive documented pleiotropy, this assumption is arguably unrealistic for genomic networks that may often have dense structures. Finally, our simulations described in detail below demonstrate that this iterative process of pruning on significance (*p* < .05 after applying an FDR multiple testing correction) closely recaptures population estimates at sample sizes typical of modern day GWAS.

### Validation via simulation

We conducted a set of realistic simulations to evaluate the performance of GNA, varying trait sample size (N = {25k, 50k, 100k, 200k, 400k}), SNP-based heritability (hsq = {0.05, 0.075, 0.10, 0.15, 0.20, 0.30}), and the number of traits in the network (K = {5, 10, 15, 20}). Each condition was repeated 100 times, such that the total number of simulation runs was 12000.

First, we generated random population networks with *K* traits, edge density of 0.4 and proportion of positive edges of 0.8, using the genGGM function in the bootnet R package. We used the GWASBrewer^73^ package to simulate GWAS summary statistics for sets of *K* traits with a given *N* and ℎ^$^, and genetically correlated as implied by the population network structure. Summary statistics were simulated for ∼1.1 million hapmap3 SNPs, with a realistic LD structure based on LD matrices calculated from 50000 random White British UK Biobank participants. The proportion of causal variants was set to 1%.

Multivariable LD score regression (Genomic SEM package) was run on each set of *K* traits to estimate the genetic covariance matrix and its associated sampling covariance matrix for each run. This is unique from standard bivariate LDSC in that it produces the noted sampling covariance matrix. GNA requires a positive definite genetic covariance matrix. Out of the 12000 simulation runs, 1206 produced non-positive definite covariance matrices. These were exclusively for low powered conditions (N <= 50k and hsq <= 0.1). Conditions with less than 70% non-positive definite runs were excluded. The genomic networks were estimated with GNA for each run with edges pruned recursively based on FDR.

### Analysing the structure of networks

#### Local network structure

Once a network has been estimated it can be further characterized using different metrics that characterize the relative importance of individual nodes within the network. These fall into two main categories of centrality metrics and clustering coefficients. Centrality metrics take various approaches to quantifying how central a node is in the network. We specifically report and describe expected influence in the main text as it may be preferred to other measures of centrality^74^ (see **Online Supplement** for description of additional measures). Expected influence quantifies the degree to which a node is directly connected to other nodes by taking the sum of the edge weights connected to that focal node. Unlike different centrality metrics that are designed to assess the node’s position in the network with distinct analytic approaches, clustering coefficients generally reflect different formulations designed to evaluate the same question of how redundant a node is in the network. This is achieved by calculating the degree to which the secondary nodes a focal node is connected to are themselves connected. A node with a high clustering coefficient thereby reflects one whose neighbours are often directly connected to one another. We specifically utilize a version of the Zhang clustering coefficient^24^ that takes into account the size and sign of the edges among neighbours^75^ and has been shown to have higher stability relative to alternative clustering coefficients^76^.

Centrality metrics and clustering coefficients can be computed on unstandardized or standardized scales. Standardized values are computed using a *Z-*score metric that is scaled relative to the mean and standard deviation for a given metric or coefficient across nodes. Values above 0 on a standardized scale thereby reflect nodes that, on average, have a higher value relative to other nodes in the network. Standardization thereby facilitates ranking and comparing nodes within the network. For this reason, the GNA package and the results presented in the main text follow the default behaviour for phenotypic network packages (e.g., qgraph^77^) to report values on a standardized scale (**Suppl. Table 5** for unstandardized results).

#### Global network structure

Insight can also be gained by analysing the properties of the overall network. The GNA R package provides three metrics to assess global network structure: 1) The global clustering coefficient (also known as transitivity) measures the overall probability for the network to have adjacent nodes interconnected^28^. It is closely related to the local clustering coefficient, but captures the degree of clustering across the entire network rather than for individual nodes. Networks with a high clustering coefficient are characteristic of random-type networks. 2) Average path length describes the average distance between nodes in the network, and is calculated as the mean of the lengths of the shortest paths between all pairs of nodes^29^. Networks with small average path lengths are characteristic of lattice-type networks. 3) The small-worldness index measures the degree to which the network possesses small-world properties^31^. Small-world networks are distinguished from random networks or lattice networks, in that they are defined as possessing *both* a high clustering coefficient and a short average path length. In GNA, we implement the small-world index proposed by Telesford and colleagues^30^, which measures small-worldness by comparing the clustering coefficient of the network, *C*, to that of an equivalent lattice network, *C_lattice_*, and comparing average path length, *L*, to that of an equivalent random network, *L_random_*: small-worldness = (*L_random_ / L*) – (*C / C_lattice_*). *C_lattice_* and *L_random_* are estimated as the mean value across 1000 simulated networks (with the same number of nodes and edges) fixed to a lattice or random structure, respectively. Values close to 0 suggest a small-world structure (*L* ≈ *L_rand_* and *C* ≈ *C_latt_*), positives values indicate the network possesses more random properties (*L* ≈ *L_rand_*, and *C* ≪ *C_latt_*), and negative values indicating more lattice network properties (*L* ≫ *L_rand_*, and *C* ≈ *C_latt_*). We note that many network indices exist beyond those described above^78^; the output from GNA can easily be used to compute these with external packages.

#### Network GWAS and TWAS

Network TWAS in GNA utilizes the covariance between imputed gene expression and each included trait. These gene expression-trait covariances reflect a rescaling of the *Z*-statistics obtained from standard univariate TWAS (e.g., those obtained from the FUSION TWAS software). For each individual gene with imputed expression, these rescaled estimates are used to expand the LDSC genetic covariance matrix to include the gene expression-trait covariances. This expanded matrix, which now includes both the gene expression-trait covariance and the trait-trait genetic covariances, is input to GNA. Additionally, the LDSC estimated sampling covariance matrix is expanded to include the sampling covariances among the TWAS estimates. GNA iteratively estimates a separate genomic network model for each gene to obtain the conditional genetic associations between the gene and the traits in the network.

In the case of a network GWAS, the LDSC estimated genetic covariance matrix is instead expanded to incorporate the covariance between the individual genetic variants and the included traits. These SNP-trait covariances reflect a rescaling of the standard univariate GWAS results. The LDSC sampling covariance matrix is expanded to incorporate the sampling covariances across the SNP-level GWAS estimates and these expanded matrices are used to then iteratively estimate the conditional genetic association between each SNP and the traits in the genomic network. The Online Supplement provides additional details about the rescaling and expansion of the genetic covariance and sampling covariance matrices needed to conduct network GWAS or TWAS. We highlight here that this rescaling is automated by existing software^9^ and the expansion of LDSC matrices is performed by the GNA R package.

An optional Ω matrix can be provided to GNA to estimate a sparse network that constrains edges between traits to be 0. This denotes that the two traits are conditionally independent of one another within the network. Which values to fix to 0 is determined by estimating the genome-wide network (i.e., not including imputed gene expression or SNP-level associations) using the default process in GNA of recursively pruning nonsignificant edges to obtain the sparse network. When using an Ω matrix, associations with gene expression or SNPs are conditional only on the included edges in the trait-trait portion of the genomic network. Taken to the extreme, if there is a singular trait with no estimated edges with other traits, then the function will reproduce the univariate TWAS or GWAS estimates for that trait. The advantage of using the Ω matrix is that conditional associations are not influenced by underpowered estimates that may increase the ratio of bias to precision. This recursively pruned Ω matrix was used for the empirical examples for psychiatric (TWAS) and metabolic/T2D (GWAS) traits.

### Empirical applications

#### Genome-wide

Prior to running GNA, all GWAS summary statistics were aligned to the same reference allele and restricted to SNPs with minor allele frequency > 1% and imputation score > 0.9 when this information was available. In addition, SNPs were restricted to HapMap3 SNPs as these tend to be well-imputed in GWAS samples and reference panel LD-scores within this subset of SNPs have been shown to produce accurate estimates of SNP-based heritability in external samples^79^. Separate LD-scores for European and East Asian genetic ancestry calculated from the 1000 Genomes Phase 3 reference data were used for LDSC estimation with GWAS of the corresponding genetic ancestry group. These LD-scores excluded the MHC region due to complex patterns of LD that can unduly bias estimates.

#### Network TWAS

For our empirical TWAS application to psychiatric disorders, univariate TWAS was run in FUSION using functional weights for the prefrontal cortex data from PsychENCODE^54^ and 13 brain tissue types from the Genotype-Tissue Expression project (GTEx v8)^53^. We utilized the quality control (QC) defaults in FUSION to restrict results to genes whose expression was imputed with an estimated accuracy of R^2^ > 0.7 and removed any genes for which > 50% of the SNPs with functional weights were missing. When restricting down to genes that passed these QC thresholds across all seven psychiatric disorders, this yielded 65,312 tissue-specific gene expression estimates for 17,297 unique transcripts. This data was then used as input to GNA to incorporate gene expression effects into the psychiatric network. Hits were defined as FDR < 0.05. FDR multiple testing correction was applied (using the *p.adjust* R package) to all p-values across both univariate and network TWAS, to ensure the FDR is controlled analysis-wide. This also carries the advantage of facilitating comparisons between disorders and between univariate and network findings, by ensuring the same significance threshold is used throughout (which would not be the case if the correction was applied per disorder). We considered the conditional association for SCZ of 12 highly-confident risk genes prioritised by the psychiatric genomics consortium in the latest SCZ GWAS, as described extensively elsewhere^45^. Briefly, these were genes supported by at least two of the following lines of evidence: i) single genes annotated to the credible set probabilistic finemapping, ii) genes strongly supported by eQTL-based Mendelian randomisation and its integration with chromatin conformation analysis (Hi-C) of adult or fetal brain, and iii) genes implicated by rare coding variants in schizophrenia, ASD or developmental disorders. We note that two of the high-confidence genes (*ZNF823* and *SLC39A8*) were not significantly associated with SCZ even in the univariate TWAS, which suggests that these risk loci are involved in SCZ through mechanisms not directly linked to mRNA expression in the bulk-tissues tested (e.g., a missense variant is the most confidently fine mapped variant in the *SLC39A8* locus^45,80^).

#### Network GWAS

The empirical network GWAS application to metabolic traits restricted the GWAS summary statistics to genetic variants with an INFO > 0.6 when this information was available. This resulted in a list of 5,753,287 SNPs that were present across the included traits. Independent loci were identified using the clumping and pruning algorithm in FUMA^81^. Significance was set at the genome-wide threshold of p < 5 x 10^-8^ and independent hits were defined as those that were not within 250 kb of one another and had an r^2^ < 0.1, where LD was defined using the 1000 Genomes East Asian Phase 3 reference sample. This same reference panel was utilized for follow-up gene set analyses (GSA) implemented with the default parameters in MAGMA^39^ that were provided either univariate GWAS or network GWAS associations with T2D as input. GSA were conducted using two different resources: 1) 17,009 gene sets and go terms defined using the Molecular Signatures Database (MSigDB)^82^, and 2) 548 gene sets defined by Anatomical Therapeutic Chemical (ATC) codes, which included all level 2, level 3 and level 4 codes for all annotated medicinal substances in DrugBank^42^. Significance for GSA results was defined at FDR < 0.05 at an FDR multiple testing correction threshold obtained from a single set of *p-*values across results obtained from univariate GWAS and network GWAS.

### Guidelines when applying GNA

#### Selecting GWAS Traits

At a minimum, we recommend only including traits with ℎ^$^ *Z*-statistic > 7 to ensure accuracy of the estimated network. This is consistent with existing recommendations for other genomic approaches utilizing LDSC data^83^ and with the indicated cut-off from our simulations for a network of five traits. If estimating a network with many traits, we recommend using our simulations as a guide for the power requirements of traits. Additionally, for traits with genetic correlations that are > 0.9 should be excluded to avoid issues with multicollinearity.

#### Evaluating the network

Provided model fit indices give an indication of how well a genomic network that is pruned on significance does of describing the data. Consistent with the field standards, we recommend a comparative fix index (CFI)^10^ > 0.9 and a standardized root mean square residual (SRMR)^11^ < 0.1. All empirical examples follow our recommended guidelines for trait selection and reported networks far exceed these model fit thresholds.

#### Run Times

Our empirical applications of trait networks (e.g., **Figures 2-4**) each took less than a minute to on a personal computer. The GNA package functions for running a network GWAS or network TWAS contain arguments that allow these processes to be run in parallel across multiple computing cores. For most applications, a network TWAS can still be run on a personal computer; our empirical application to seven psychiatric disorders and 65,312 imputed gene expression levels took 28.1 minutes when run in parallel on a personal computer with 8 cores. For network GWAS applications, and similar to traditional GWAS, it will be pragmatic to perform analyses on a computing cluster environment. The conditional associations estimated for the genes and genetic variants in GNA are all independent of one another, which further allows for splitting analyses across multiple jobs without affecting results. Our empirical network GWAS application to six traits (T2D and cardiometabolic traits) for 5,753,287 SNPs was split across 10 jobs on computing nodes with 36 cores each, wherein each job finished in an average of 38.6 minutes. Collectively, these run times indicate that GNA analyses are computationally efficient and can be expediently completed on a personal computer for trait network and network TWAS applications and on a computing cluster for network GWAS applications.

## Data Availability

The data used in this study are all publicly available or can be requested for access. Specific download links for various datasets are directly below.
Summary statistics for cardiometabolic traits in East Asian ancestry are available to download on GWAS catalog: https://www.ebi.ac.uk/gwas/
Psychiatric disorder summary statistics for data from the psychiatric genomics consortium (PGC) can be downloaded here: https://www.med.unc.edu/pgc/download-results/
Links to the GTEx v8 gene expression reference weights used for univariate TWAS imputation in FUSION can be found here: http://gusevlab.org/projects/fusion/
Links to the functional reference weights from PsychENCODE also used for univariate TWAS can be found here: http://resource.psychencode.org/

https://www.med.unc.edu/pgc/download-results/

http://gusevlab.org/projects/fusion/

http://resource.psychencode.org/

## Acknowledgements

JGT is supported by an Australian National Health and Medical Research Council (NHMRC) EL1 Investigator Grant (2027002). WRR is supported by an Australian NHMRC EL1 Investigator Grant (2025671). EMD is supported by an Australian NHMRC L1 Investigator Grant (2026364). ADG is supported by NIH Grants R01MH120219 and RF1AG073593.

## Author Contributions

*Methods development:* J.G.T., A.D.G.

*Software development:* J.G.T., A.D.G.

*Empirical applications:* J.G.T., Z.F.G., W.R.R., E.M.D., A.D.G

*Writing*: J.G.T., W.R.R., A.D.G.

*Feedback and editing:* J.G.T., Z.F.G., W.R.R., E.M.D., A.D.G.

