## Supplementary Information for "Genomic network analysis characterizes genetic architecture and identifies trait-specific biology"

### Additional centrality metrics

*Strength.* Strength is a measure of how strongly a node is directly connected and is calculated as the sum of the absolute value of the edge weights for a given node.[1] We note that strength and expected influence (detailed in **Method** of main text) will produce the same values if all edges are either positive or negative.[2] Like expected influence, it can be considered a measure of overall direct influence (i.e., does not consider mediating role of other nodes). Expected influence will likely be preferred when the signs of the edges have an inherent meaning, as will generally be the case in GNA. For example, for our empirical application to 12 neuroticism items we are interested not just in which nodes are connected irrespective of directionality, but rather which nodes have conditionally positive genetic associations with other items.

*Closeness and Betweenness.* While strength and expected influence evaluate the direct influence of a focal node, closeness and betweenness consider indirect connections to other nodes via the focal node. Closeness measures the distance between a focal node and the remaining nodes. It is calculated as the sum of the inverse distances for the focal node with all other nodes in the network.[3] Betweenness assesses the degree to which the shortest paths between two nodes in the network are connected via the focal node[4]. Consequently, removing a node with larger betweenness will generally result in longer distances between the remaining nodes[5]. Conversely, if a node with high betweenness is changed, that change is expected to more quickly propagate throughout the rest of the network. Betweenness and closeness are calculated using distances that do not take into account the size of the edge weights[6], which has been highlighted as problematic[7]. These metrics arguably provides a less nuanced way of examining differentiated networks in GNA where edge weights may evince substantial variability across the genomic network.

*Empirical Neuroticism Example.* Evaluating node importance in this sparse network revealed that nervousness had the strongest expected influence and, relative to other nodes in the network, higher betweenness and centrality and a lower clustering coefficient. This indicates that the genetic component of nervousness reflects a crucial component that uniquely links various aspects of neuroticism (**Suppl. Fig. 1).**

### Graph options

Once a network is estimated, the results can be visualized using several different graphing approaches. Below we briefly consider the benefits and limitations of five frequently utilized graphing options in the network literature that can be requested as output from GNA in the open-source *GNA* R package. A more complete explanation of the statistical properties, benefits, and limitations of each visualizing option is also provided in an overview paper from Jones et al. (2018)[8].

***Spring Graphs*****.** *Benefits:* Spring graphs are based on force-directed algorithms (most commonly the Fruchterman-Reingold[9]), which produce nodes that rarely visually overlap along with edges that are evenly spaced. The corresponding benefits are visually appealing graphs with easier to view relationships. *Limitations:* The primary limitation is that the positioning of nodes is not interpretable, meaning spatial proximity between nodes does not indicate strength of connection. This can lead to misinterpretation of node closeness as reflecting similarity. For this reason, it is important for authors to clarify what type of graphing option was used to avoid overinterpretation of the visual representation from spring graphs.

***Circle Graphs.*** *Benefits:* As the name implies, circle graphs place all nodes in a circle. These graphs are aesthetically simple and easy to interpret, especially when the emphasis is on equalizing node spacing and providing a clear, symmetrical layout for visual inspection. In addition, the circle graph lends itself to direct comparisons across different networks of associations applied to the same set of traits. For example, we use the circle graph for two of our empirical examples (for 12 neuroticism items and type 2 diabetes with other cardiometabolic traits) to highlight how the multivariate structure becomes increasingly simplified when going from genetic correlations to the pruned set of significant, partial genetic correlations produced from GNA. *Limitations:* Similar to spring graphs, the spatial positioning does not provide meaningful information about the relationships between nodes and no inferences should be drawn about node distances.

***Multidimensional Scaling (MDS).*** *Benefits:* MDS graphs function to reduce high-dimensional data down to a two-dimensional space that is readily visualized[10]. This two-dimensional space in MDS is referred to as the configuration matrix, which reflect the Cartesian coordinates for plotting the network model. These coordinates are chosen so that the distance between nodes is an approximate function of the edge weight between a pair of nodes. Thus, the placement of nodes on the graph is generally meaningful when using MDS visualization. *Limitations:* There are times when the two-dimensional solution may not be able to capture the complexity of the network, and in these cases the proximity of nodes to one another may not accurately depict node similarity[11].

***PCA Graphs****. Benefits:* Graphs in GNA based on principal components analysis (PCA) plot the eigenvalue decomposition applied to the genetic correlation matrix on the first two principal components along the X and Y axis. The distance between nodes along each of these axes is therefore interpretable and provides information on their relationship along these two latent dimensions. In certain use cases, these dimensions may have additional interpretive value if there is theoretical reasons or prior analyses that indicate the nodes in the network cluster along conceptually similar dimensions. As with MDS, the PCA plotting method can help simplify complex data and present relationships along interpretable axes. *Limitations:* Also like MDS, two-dimensional solutions may not be sufficient for describing the complexity of a given network and underlying patterns in the data captured beyond the first two PCs may be lost. In addition, if nodes strongly cluster together along a particular dimension, this can render visual interpretation difficult as the edges connecting these nodes will be overlapping and hard to discern from one another[8]. This is the primary reason that PCA graphs were not used for the empirical examples in the current paper (i.e., clustering of nodes along the two dimensions produced a difficult to parse visual representation of the network structure).

***Eigenmodel Graphs.*** *Benefits:* Similar to PCA, eigenmodel graphs plot the eigenvalues along the first two dimensions along the X and Y axis. Unlike PCA which is applied to the genetic correlation matrix, the eigenmodel graph technique extracts latent dimensions from the structure of the network itself (i.e., the partial genetic correlations in GNA). As a result, it can represent relational patterns and clusters in the network, allowing researchers to explore hidden structures, such as the types of node communities indexed by global metrics (described in **Method** section of main text). *Limitations:* The eigenmodel graphs contain the same limitation of PCA that visualization can be difficult when certain traits cluster strongly together along either of the two dimensions. In addition, latent dimensions estimated from partial correlations (genetic or phenotypic) are less common in the literature, making it more difficult to map results from this method onto previously described constructs.

**Model fit**

Model fit statistics are typically calculated using participant sample *(N).* However, our genomic network approach allows for including GWAS summary statistics that will often vary in contributing sample size. To circumvent this issue, the *GNA* package calculates several commonly used model fit statistics using the sampling covariance matrix (*V*) that describes the range in precision across the included GWAS. These model fit statistics can then be used to evaluate whether more parsimonious (i.e., sparse) networks that have been pruned at some threshold continue to adequately describe the observed genetic covariance matrix. Below we describe how these different metrics are calculated.

Model $\chi^2$ reflects an aggregate index of the discrepancy between the model implied and observed covariance matrix. Our equation starts by calculating the residual covariance matrix (*R*) as:

$$R = S - \sum(\theta) \quad (1)$$

where *S* is the observed covariance matrix and $\sum(\theta)$ reflects the model implied covariance matrix from the network model. We then weight this residual matrix by the precision of the cells within that matrix by first taking the eigendecomposition of the sampling covariance matrix (*V*):

$$V = (\mathrm{P}_1\ \mathrm{P}_0)\begin{pmatrix} E & 0 \\ 0 & 0 \end{pmatrix}\begin{pmatrix} P_1' \\ P_o' \end{pmatrix} \quad (2)$$

where $P_1$ is the matrix of principal components (eigenvectors) of *V*, $P_0$ is the null space of *V*, and $E$ is a diagonal matrix of the non-zero eigenvalues of *V*. These eigenvalues and eigenvectors can then be used to weight the residual covariance matrix to obtain a $\chi^2$ distributed test statistic given as:

$$\chi^2(df) \sim R_i'\mathrm{P}_1 E^{-1}\mathrm{P}_1' R_i \quad (3)$$

where *df* reflects the degrees of freedom, calculated as the difference between the number of unique elements in the *S* matrix and the number of freely estimated edges in the network model, and $R_i$ is the residual matrix in vectorized form.

The Comparative Fit Index (CFI) provides an alternative index of model fit, so named as it compares the fit of the proposed model to the fit of what is referred to as the independence model. This independence model reflects one in which only the variances of the traits are modelled (i.e., traits are specified to be entirely independent of one another). CFI is specifically calculated as:

$$\frac{f(\text{Independence Model}) - f(\text{Proposed Network Model})}{f(\text{Independence Model})} \quad (4)$$

where $f$ is calculated for both the independence and network model as: $\chi^2 - df$. The $\chi^2$ of the independence model is then calculated as above, with the exception that the vector of residuals, $R_i$, is taken from a residual matrix that is simply calculated by setting only the diagonal elements of the genetic covariance matrix to 0:

$$R_{independence} = S - diag(diag(S)) \quad (5)$$

We note that the formulas for model $\chi^2$ and CFI follow formulations described previously[12], but improve upon these prior specifications by removing the need to estimate any follow-up models, thereby guaranteeing more stable and computationally efficient solutions.

Akaike Information Criterion (AIC) is a fix index that counterbalances the misfit of the model, as indexed by model $\chi^2$, by subtracting out two times the degrees of freedom of the specified network:

$$\text{AIC} = \chi^2 - 2df \quad (6)$$

AIC can be used to compare network models even when they are not nested. Standardized Room Mean Square Residual (SRMR) is the one model fit metric that we provide that does not traditionally rely on $N$ and can be calculated as usual. This reflects the square root of the average squared difference between the model-implied correlation matrix and the observed correlation matrix (i.e., on a standardized scale).

**Network TWAS**

GNA takes as input the results from LDSC and the output from univariate TWAS across all included traits. This allows GNA to expand the genetic covariance matrix to include the covariances between imputed expression for a given gene, $j$, and the included traits (1 through $k$):

$$\begin{bmatrix} \sigma^2_{Gene_j} & & & & & \\ \sigma_{Gene_j,1} & h^2_1 & & & & \\ \sigma_{Gene_j,2} & \sigma_{1,2} & h^2_2 & & & \\ \sigma_{Gene_j,3} & \sigma_{1,3} & \sigma_{2,3} & h^2_3 & & \\ \vdots & \vdots & \vdots & \vdots & \ddots & \\ \sigma_{Gene_j,k} & \sigma_{1,k} & \sigma_{2,k} & \sigma_{3,k} & \cdots & h^2_k \end{bmatrix} \quad (7)$$

The value in the first cell of the matrix, $\sigma^2_{Gene_j}$, reflects the heritability of the expression of an individual gene captured by cis-effects, which is provided directly by the FUSION

TWAS software[13]. The remaining values are the LDSC genetic covariance matrix (which we call *S*), with the $h_k^2$ values on the diagonal reflecting the SNP-based heritabilities and the $\sigma_{k,k}$ values the genetic covariances between each pairwise combination of traits. GNA currently uses listwise deletion wherein only genes with estimated effects that pass QC thresholds across all traits can be incorporated into the genomic network.

To appropriately add the gene expression effects to this expanded matrix, the results from TWAS must first be rescaled to appropriately mirror the scaling of the LDSC estimates in the *S* matrix. More specifically, the elements of the *S* matrix reflect heritability and covariance estimates that are partially standardized. We use the term partially standardized to refer to the fact that they are standardized relative to the total variance in the phenotype (i.e., the GWAS trait), but not standardized relative to the genetic predictors (i.e., the genotypes). In FUSION, TWAS results are provided as *Z*-statistics, which must be turned into a partially standardized regression coefficient and its corresponding *SE*. As has been described previously[14], for continuous traits this can be calculated as $b_{Gene_j,k} = \frac{TWAS\ Z_{k,j}}{\sqrt{N\sigma^2_{Gene_j}}}$ and $SE_{b_{Gene_j,k}}$ = $\frac{b_{Gene_j,k}}{TWAS\ Z_{k,j}}$. For binary traits (e.g., case/control GWAS outcomes), the TWAS *Z*-statistics are converted to an unstandardized logistic regression coefficient and its *SE*. This can be calculated as $blogit^*_{Gene_j,k} = \frac{TWAS\ Z_{k,j}}{\sqrt{\sum n_i v_i (1-v_i) n_i\, \sigma^2_{Gene,j}}}$. The term in the denominator $\sum n_i v_i (1 - v_i)$ reflects the sum of the sample sizes across the cohorts contributing to the univariate GWAS meta-analysis, with a correction to the sample size within each cohort, *i,* that do not have a balanced proportion of 50% cases and 50% controls, where $v_i$ and $n_i$ reflect the cohort-specific sample prevalence and sample size, respectively[15]. The corresponding *SE* is calculated as $SE_{\ blogit^*_{Gene_j,k}} = \frac{blogit^*_{Gene_j,k}}{TWAS\ Z_{k,j}}$ .This unstandardized logistic regression coefficient and its *SE* are then divided by variance of the phenotype on the liability scale, which is given as: $\sqrt{\sigma^2_{Gene_j} \times blogit^2_{Gene_j,k} + \frac{\pi^2}{3}}$, where $\sigma^2_{Gene_j} \times blogit^2_{Gene_j,k}$ reflects the variance in the outcome, *k,* predicted by the gene, *j,* and $\frac{\pi^2}{3}$ is the residual variance from a logistic regression. Finally, both continuous and logistic coefficients are converted to covariance estimates by taking the product of these coefficients and the variance of the gene ($\sigma^2_{Gene_j}$) as $\sigma_{Gene_j,k}$= $b_{Gene_j,k} \times \sigma^2_{Gene_j}$ and $SE_{\sigma_{Gene_j,k}} = SE_{b_{,gk}} \times \sigma^2_{Gene_j} \times \sqrt{Na+1}$. The $Na + 1$ term reflects the unviariate intercept from LDSC that consists of *N,* the sample size for the trait, and *a,* the uncontrolled population stratification from the GWAS. Including this term serves to correct the standard errors for uncontrolled stratification. No correction on the *SEs* is applied when the univariate LDSC intercepts are < 1.

With the genetic covariance matrix (*S*) expanded to included trait-gene expression covariances, the corresponding sampling covariance matrix ($V_s$) must also be expanded. The portion of the sampling covariance matrix that indexes the gene expression effects contains the squared *SE*s of the gene-phenotype covariances on the diagonal. The sampling dependencies between these TWAS estimates on the off-diagonal is indexed via the unstandardized bivariate LDSC intercept. The bivariate intercept is estimated directly from the GWAS data when running multivariable LDSC and can be expressed as: $\sqrt{N_1 N_2}a + \frac{\rho N_s}{\sqrt{N_1 N_2}}$, where the *Ns* are the sample sizes of the two GWAS, *a* is shared residual population stratification not controlled for in both univariate GWAS, and $\rho$ is the phenotypic correlation among overlapping participants. As the bivariate intercept is a sampling correlation, this is rescaled to a sampling covariance (i.e., unstandardized) using the squared *SEs* (sampling variances) of the gene-genotype covariances that populate the diagonal of this portion of $V_s$. The sampling covariance between the gene-genotype covariances and the genome-wide estimates from LDSC ($h^2_{SNP}$ and genetic covariances) are fixed to 0. This is a realistic simplifying assumption that reflects the fact that imputed gene expression will be approximately independent of any other genetic effects except for those in shared LD blocks. GNA automatically expands the *S* and $V_s$ matrices for the user for each individual gene prior to estimating a network that incorporates that gene.

**Network GWAS**

When running a network GWAS, GNA expands the genetic covariance matrix to include the SNP-phenotype covariances for each SNP, *j,* and the included traits (1 through *k):*

$$\begin{bmatrix} \sigma^2_{SNP_j} & & & & & \\ \sigma_{SNP_j,1} & h^2_1 & & & & \\ \sigma_{SNP_j,2} & \sigma_{1,2} & h^2_2 & & & \\ \sigma_{SNP_j,3} & \sigma_{1,3} & \sigma_{2,3} & h^2_3 & & \\ \vdots & \vdots & \vdots & \vdots & \ddots & \\ \sigma_{SNP_j,k} & \sigma_{1,k} & \sigma_{2,k} & \sigma_{3,k} & \cdots & h^2_k \end{bmatrix} \quad (8)$$

Ordinary least squares (OLS) or logistic regression coefficients provided for GWAS summary statistics of quantitative or binary traits, respectively, are transformed into SNP-phenotype covariances, and corresponding standard errors, in two steps. First, GWAS results are rescaled to reflect partially standardized regression coefficients and standard errors that are standardized with respect to the variance in the trait, but not the variance in the genotype. For quantitative traits, these coefficients are calculated from GWAS *Z*-statistic (calculated as the ratio of the GWAS estimate over its standard error) as: $b_{SNP_j,k} = \frac{GWAS\ Z_{k,j}}{\sqrt{N\sigma^2_{SNP_j}}}$ and $SE_{b_{SNP_j,k}} = \frac{b_{SNP_j,k}}{GWAS\ Z_{k,j}}$ , where $\sigma^2_{SNP_j}$ reflects the variance of a given SNP, *j*. $\sigma^2_{SNP_j}$ is calculated as *2pq*, where *p* = the minor allele frequency (MAF) and *q* = 1-MAF. These MAFs can be taken directly from the GWAS summary statistics or from a reference sample (e.g., 1000 Genomes) when this information is not available. We have reassuringly demonstrated in prior work that using reference sample versus in-sample MAFs produces highly concordant sets of results[12]. For binary traits, the logistic regression coefficient and its *SE* are divided by the variance of the phenotype on the liability scale, which is given as: $\sqrt{\sigma^2_{SNP_j} \times blogit^2_{SNP_{j},k} + \frac{\pi^2}{3}}$. Second, the partially standardized regression coefficients are rescaled to reflect SNP-phenotype covariances as $\sigma_{SNP_{j},k} = b_{SNP_{j},k} \times \sigma^2_{SNP_j}$ and $SE_{\sigma_{SNP_{j},k}} = SE_{b_{SNP_{j},k}} \times \sigma^2_{SNP,j} \times \sqrt{Na + 1}$. The univariate LDSC intercept $(Na + 1)$ serves to correct *SE*s for uncontrolled population stratification and no correction is applied when the intercept is < 1. The sampling covariance matrix, $V_s$, is expanded mirroring the process described above for TWAS, which indexes the sampling covariances across SNP-phenotype estimates for two traits using the unstandardized cross-trait LDSC intercept.

A specific transformation is applied when either only GWAS *Z*-statistics are available for binary traits or a linear proability model has been used where an OLS GWAS is used for a binary outcome . A linear probability model is applied for pragmatic reasons as OLS regression is typically much faster computationally than its logistic counterpart. This approach is justified by the fact that both approaches tend to produce highly concordant sets of *Z*-statistics when the effect sizes for the predictors (the individual SNPs) are small[16], with the main distinction in the results between a linear probability model and logistic regression being the scaling of the GWAS effect estimates and standard errors. An approximation of the logistic regression coefficient can be calculated from the GWAS *Z*-statistics using a similar equation used to back out logistic betas from TWAS *Z*-statistics. For GWAS *Z*-statistics, these are calculated as $blogit^*_{SNP_{j},k} = \frac{GWAS\ Z_{k,j}}{\sqrt{\Sigma n_i v_i (1 - v_i) n_i \sigma^2_{SNP,j}}}$ with the corresponding *SE* given as $SE_{blogit^*_{SNP_{j},k}} = \frac{blogit^*_{SNP_{j},k}}{GWAS\ Z_{k,j}}$. Mirroring the procedure described above for instances where logistic regression coefficients are directly avialbale in the GWAS summary statistics, these estimates are then rescaled relative to the variance of the phenotype on the liability scale and finally transformed into SNP-phenotype covariances.

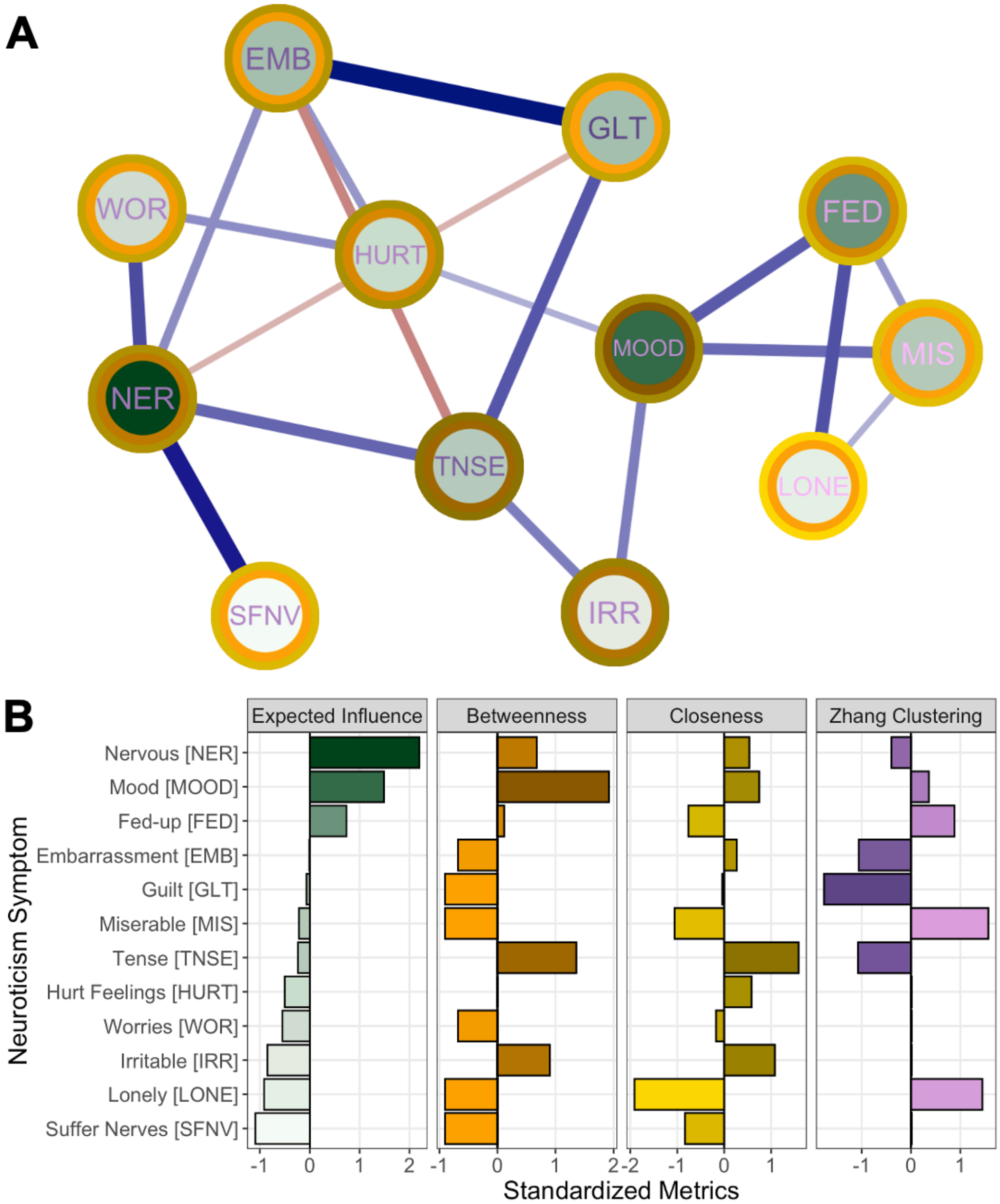

**Supplementary Figure 1. Centrality and Clustering in Neuroticism Network.** *Panel A.* The neuroticism network plotted using multidimensional scaling. The circles are shaded according to their expected influence, the inner border shaded according to betweenness, the outer border shaded by closeness, and the text shaded according to the clustering coefficient. *Panel B.* Centrality (expected influence, betweenness, and closeness) metrics and the Zhang clustering coefficient[17]. For comparison, all values are ordered according to the level of expected influences and all values were standardized relative to the mean and standard deviation of that metric's values across nodes. Abbreviations for neuroticism items are defined in the *y*-axis of *Panel B.* For both panels darker shading indicates larger and smaller values on the three centrality metrics and Zhang clustering coefficient, respectively, such that darker shading across metrics indicates a more critical node in the network.

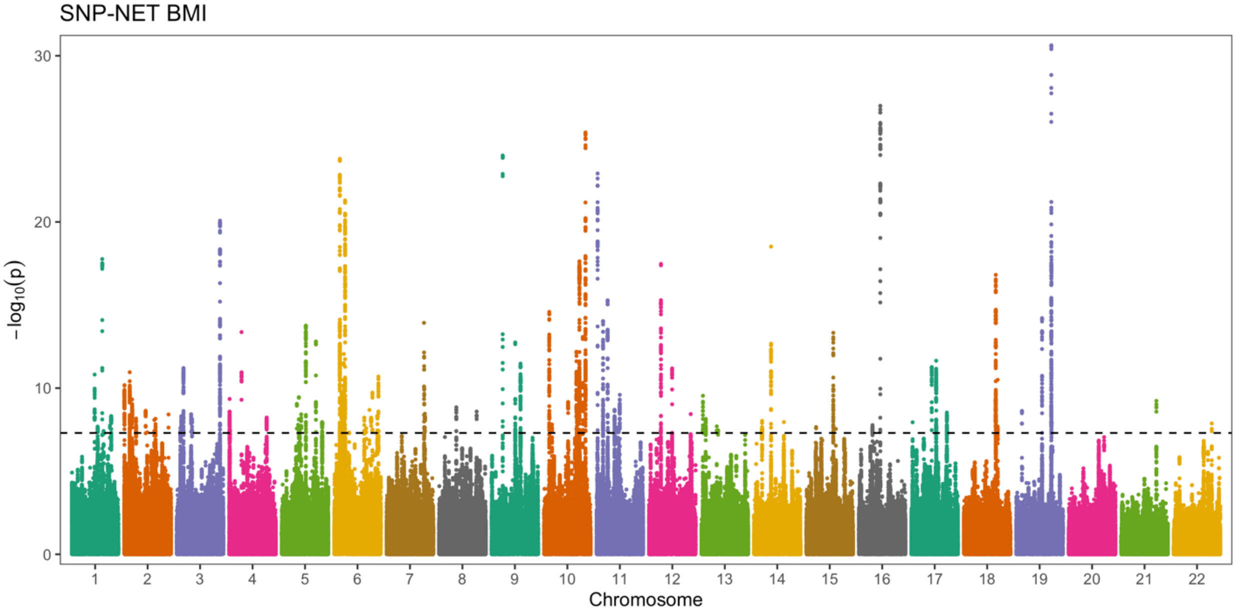

**Supplementary Figure 2. Body Mass Index Network GWAS Manhattan Plot.** Manhattan plot depicts network GWAS results for body mass index (BMI). Results reflect associations between genetic variants (SNPs) and the unique genetic variance in BMI controlling for the other estimated edges in the final network (depicted in **Figure 1** of main text). The dashed line indicates genome-wide significance ($p$ < 5e-8). A list of independent loci can be found in Supplementary Table 11.

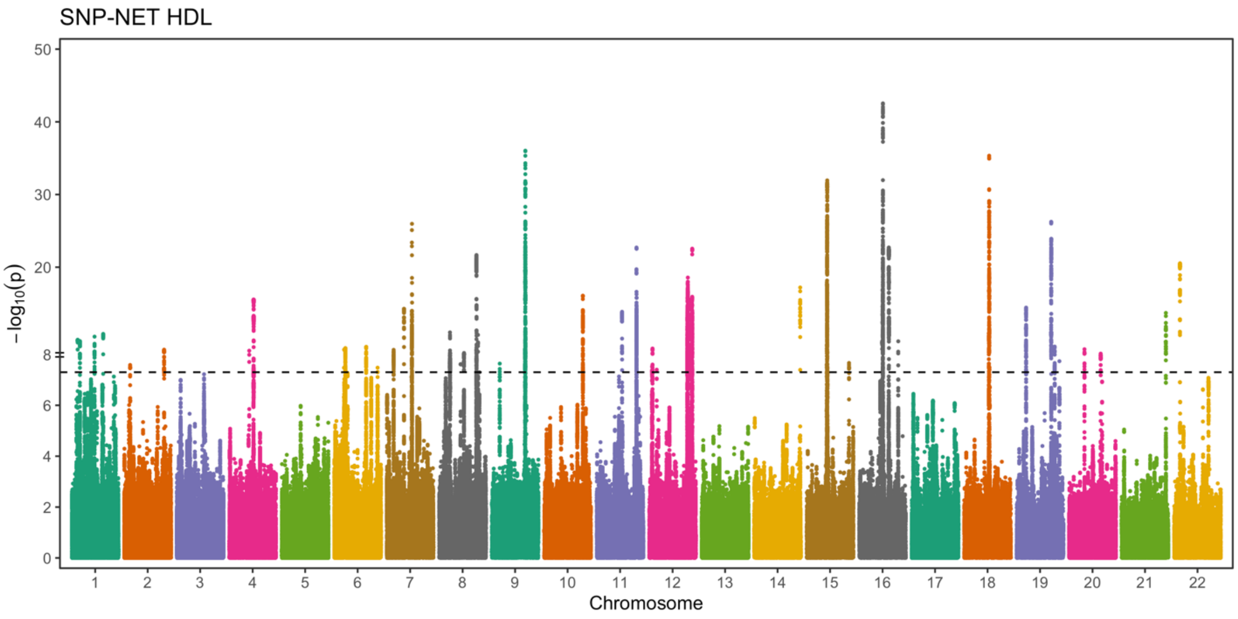

**Supplementary Figure 3. High Density Lipoprotein Network GWAS Manhattan Plot.** Manhattan plot depicts network GWAS results for high density lipoprotein (HDL). Results reflect associations between genetic variants (SNPs) and the unique genetic variance in HDL controlling for the other estimated edges in the final network (depicted in **Figure 1** of main text). The dashed line indicates genome-wide significance ($p$ < 5e-8). A list of independent loci can be found in Supplementary Table 13.

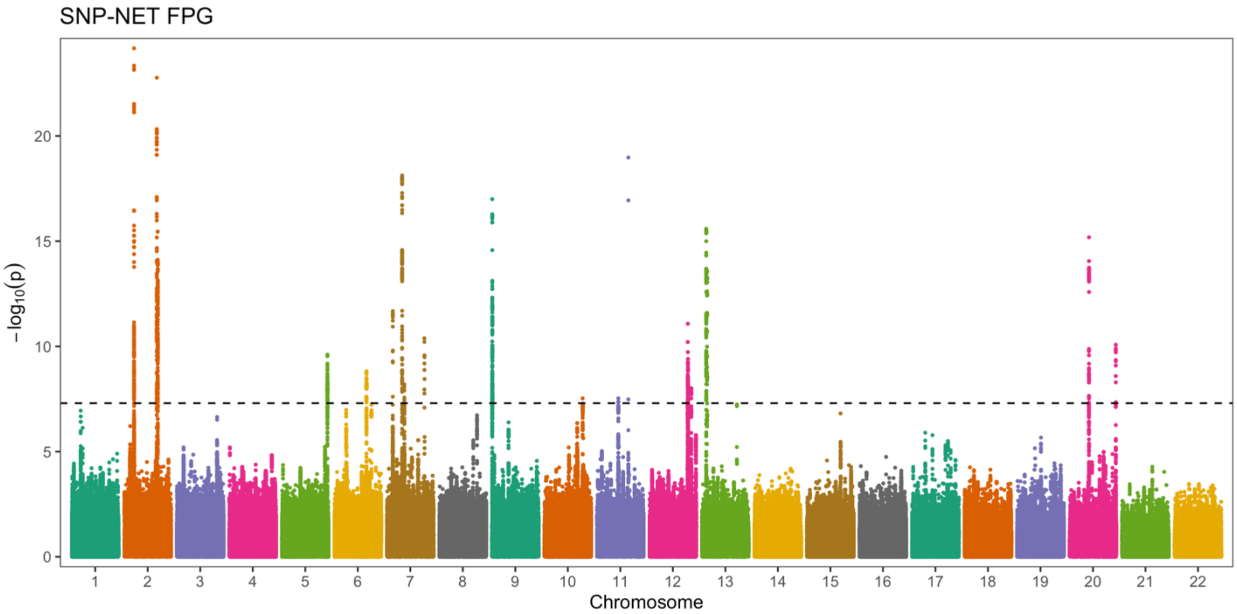

**Supplementary Figure 4. Fasting Plasma Glucose Network GWAS Manhattan Plot.** Manhattan plot depicts network GWAS results for fasting plasma glucose (FPG). Results reflect associations between genetic variants (SNPs) and the unique genetic variance in FPG controlling for the other estimated edges in the final network (depicted in **Figure 1** of main text). The dashed line indicates genome-wide significance ($p$ < 5e-8). A list of independent loci can be found in Supplementary Table 12.

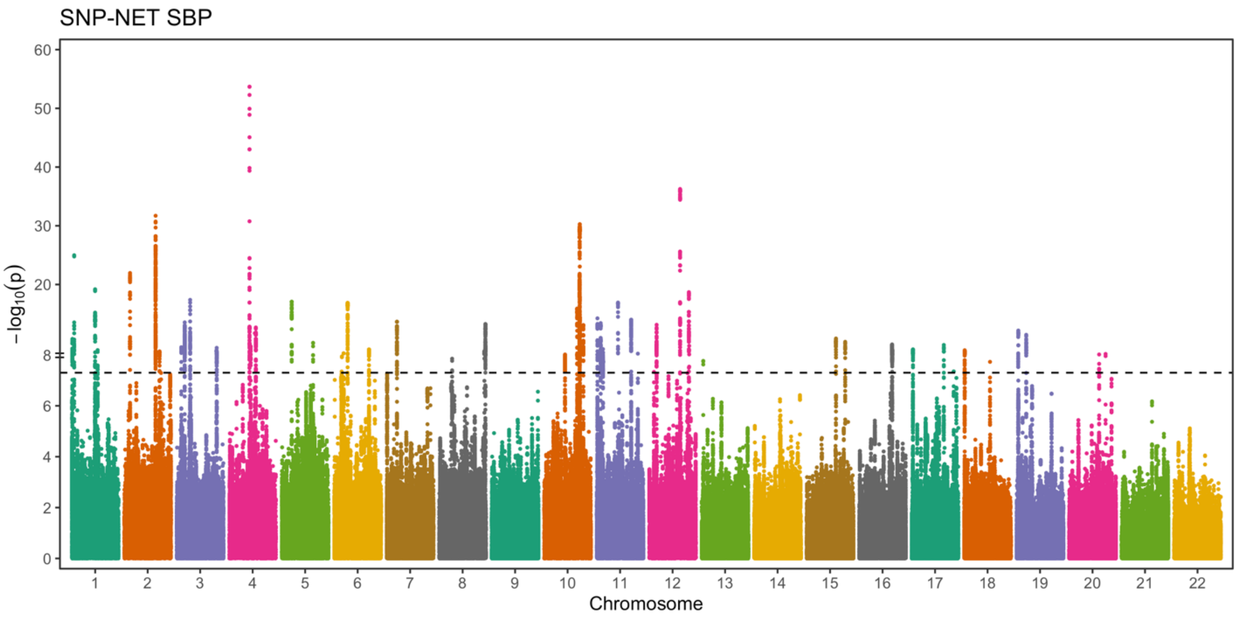

**Supplementary Figure 5. Systolic Blood Pressure Network GWAS Manhattan Plot.** Manhattan plot depicts network GWAS results for systolic blood pressure (SBP). Results reflect associations between genetic variants (SNPs) and the unique genetic variance in SBP controlling for the other estimated edges in the final network (depicted in **Figure 1** of main text). The dashed line indicates genome-wide significance ($p$ < 5e-8). A list of independent loci can be found in Supplementary Table 14.

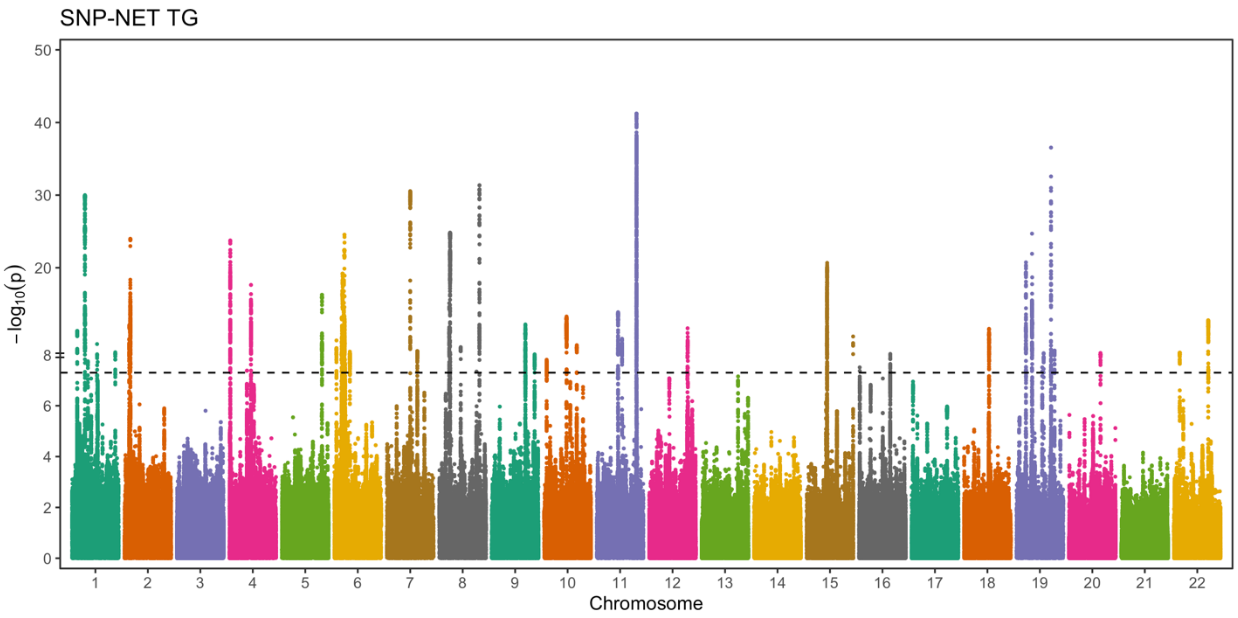

**Supplementary Figure 6. Triglycerides Network GWAS Manhattan Plot.** Manhattan plot depicts network GWAS results for triglycerides (TG). Results reflect associations between genetic variants (SNPs) and the unique genetic variance in TG controlling for the other estimated edges in the final network (depicted in **Figure 1** of main text). The dashed line indicates genome-wide significance (*p* < 5e-8). A list of independent loci can be found in Supplementary Table 15.
